# Optimizing antibiotic use in Indonesia: a systematic review and synthesis of current evidence to inform opportunities for intervention

**DOI:** 10.1101/2022.02.20.22271261

**Authors:** Ralalicia Limato, Gilbert Lazarus, Puck Dernison, Manzilina Mudia, Monik Alamanda, Erni J. Nelwan, Robert Sinto, Anis Karuniawati, H. Rogier van Doorn, Raph L. Hamers

## Abstract

**Introduction:** A major driver of antimicrobial resistance (AMR) and poor clinical outcomes is suboptimal antibiotic use, although data are lacking in low-resource settings. We reviewed studies on systemic antibiotic use (WHO ATC/DDD category J01) for human health in Indonesia, and synthesized available evidence to identify opportunities for intervention.

**Methods:** We systematically searched five international and national databases for eligible peer-reviewed articles, in English and Indonesian, published between 1 January 2000 and 1 June 2021 including: 1) antibiotic consumption; 2) prescribing appropriateness; 3) antimicrobial stewardship (AMS); 4) perceptions among consumers and providers. Two independent reviewers included studies and extracted data. Study-level data were summarized using random-effects model meta-analysis for consumption and prescribing appropriateness, effect direction analysis for AMS interventions, and qualitative synthesis for perception surveys. (PROSPERO CRD42019134641)

**Results:** Of 9323 search hits, we included 100 reports on antibiotic consumption (20), prescribing appropriateness (49), AMS (13), and/or perception (25) (8 categorized in >1 domain). The pooled estimate of overall antibiotic consumption was 110.1 DDD/100 patient-days (95%CI98.5-121.6), with ceftriaxone, ampicillin and levofloxacin being most consumed. Pooled estimates for overall appropriate prescribing (according to Gyssens method) were 33.5% (95%CI18.1-53.4%) in hospitals and 49.4% (95%CI23.7-75.4%) in primary care. Pooled estimates for appropriate prescribing (according to reference guidelines) were, in hospitals, 99.7% (95%CI97.4-100%) for indication, 84.9% (95%CI38.5-98.0%) for drug choice, and 6.1% (95%CI0.2-63.2%) for overall appropriateness, and, in primary care, 98.9% (95%CI60.9-100%) for indication, 82.6% (95%CI50.5%-95.7%) for drug choice and 10.5% (95%CI0.8-62.6%) for overall appropriateness. The few AMS intervention studies conducted to date suggested potential to reduce antibiotic consumption and improve prescribing appropriateness. Key themes identified in perception surveys were lack of antibiotic knowledge among consumers and non-prescription antibiotic self-medication.

**Conclusions:** Context-specific strategies are urgently needed to improve rational antibiotic use in Indonesian hospitals and communities, with critical evidence gaps concerning private and informal health providers.

**KEY QUESTIONS:** **What is already known?**

• Indonesia is a potential AMR hotspot, where, based on pharmaceutical sales data, antibiotic consumption increased 2.5-fold between 2000 and 2015, mostly driven by broad-spectrum penicillins, fluoroquinolones and cephalosporins.
• Representative contemporary data on antibiotic use are lacking, although anecdotal data suggest antibiotic overuse in the healthcare system, widespread over-the-counter use in communities, and high rates of AMR mostly among common Gram-negative bacteria.
• A comprehensive review on antibiotic use in human health in Indonesia has not been conducted to date.

**What are the new findings?**

• Available data spanning the past 20 years, suggested that only 34% and 49% of antibiotics were appropriately prescribed in hospital and primary care settings, respectively, although the quality of the evidence was low.
• Publications evaluating AMS interventions have been sparse to date, demonstrating the need to strengthen the local research base to develop context-specific and sustainable AMS models.
• Community surveys suggested important gaps in antibiotic knowledge, and that non-prescription antibiotic self-medication is common practice, although data to quantify this problem and its drivers are lacking.

**What do the new findings imply?**

• Available evidence synthesised in this Review provides important insights in the magnitude and patterns of antibiotic use, and associated patient and health system factors, which helps define opportunities for optimising responsible antibiotic use.
• Critical evidence gaps exist on informal and formal private health care providers, geographic areas outside of Java Island, as well as effective AMS models that consider country-specific socio-cultural, economic and political circumstances.
• Optimization of antimicrobial use as a means to tackle AMR should be a priority of the national agenda for universal health coverage.

## INTRODUCTION

The global rise in antimicrobial resistance (AMR) is considered one of the greatest public health threats worldwide, with a disproportionate impact in low- and middle-income countries (LMIC).^1^ A recent global analysis estimated that AMR was directly responsible for 1.27 million deaths and played a part in 4.95 million deaths in 2019.^1^ One of the major drivers of antimicrobial resistance (AMR) is antibiotic use, including their overuse and misuse, for human health.^2, 3^ In low-resource settings, lack of access to quality health care, vaccination, safe water and sanitation, leave many people vulnerable to infection and dependent on antibiotics for treatment, with their use largely unregulated. At the same time, access to life-saving drugs targeting the emerging resistant pathogens remains an issue in many settings. Globally, during the past decade concerted efforts have been made to develop strategies to preserve the effectiveness of existing antibiotic agents, although this has been partially set back because of the COVID-19 pandemic.^4^

Indonesia is a lower-middle-income country in Southeast Asia with the world’s fourth largest population (274 million), and socio-economic conditions and health indicators vary widely across the archipelago. More than 55% of the population is concentrated on Java Island, which contributes around 59% to the national economy and has the best developed health infrastructure.^5^ Based on pharmaceutical sales data between 2000 and 2015, Indonesia ranks among the greatest risers in antibiotic consumption (29^th^ of 76 countries).^3^ A range of complex factors, including variable access to quality health care, persistently high infectious disease burdens,^6^ and weakly enforced antibiotic policies, render Indonesia particularly vulnerable to AMR.^5–7^ The implementation of the National Action Plan for AMR, launched in 2017^8, 9^ has been hindered due to, among other factors, a limited evidence base of AMR epidemiology, antibiotic utilisation and rational prescribing practices.^7^

We systematically reviewed the scientific literature on antibiotic use for human health, in both hospital and primary care settings, in Indonesia spanning the past 20 years. This Review focused on four key domains: 1) antibiotic consumption; 2) appropriateness of antibiotic prescribing; 3) AMS interventions; 4) knowledge, attitudes and perceptions among consumers and providers. This Review also reflects on current progress in the National Action Plan for AMR, and defines evidence gaps and context-specific priorities for action.

## METHODS

### Search strategy and selection criteria

This systematic review was reported according to the Preferred Reporting Items for Systematic Reviews and Meta-analyses (PRISMA) 2020 guidelines.^10^ A detailed protocol has been prospectively registered in the PROSPERO database (CRD42019134641). Data for this systematic review were identified through the screening of five international (PubMed, EMBASE and Google Scholar) and national (Garuda [Garba Rujukan Digital] and Neliti) electronic bibliographic databases. Reference screening was used to identify additional relevant papers. The review was limited to antibacterials for systematic human use (J01 category in WHO ATC/DDD index). Only peer-reviewed original articles published in English or Indonesian between 1 January 2000 and 1 June 2021 were included. Search terms covered all domains of this Review and combined Indonesia, antibiotic, antimicrobial, consumption, prescription, dispensing and stewardship. Full search terms are listed in **Table S1**. We did not contact study authors.

**Table 1.**
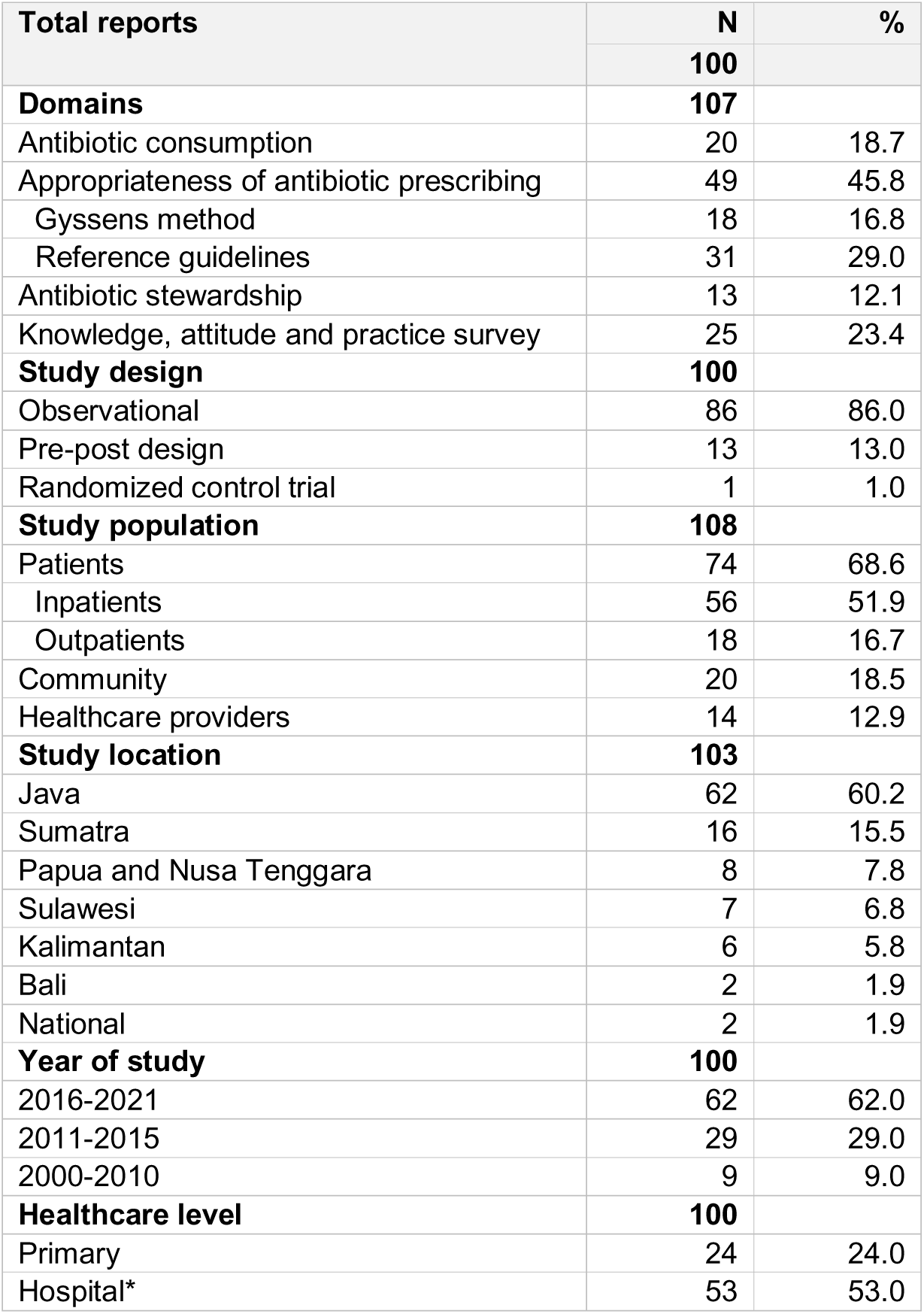
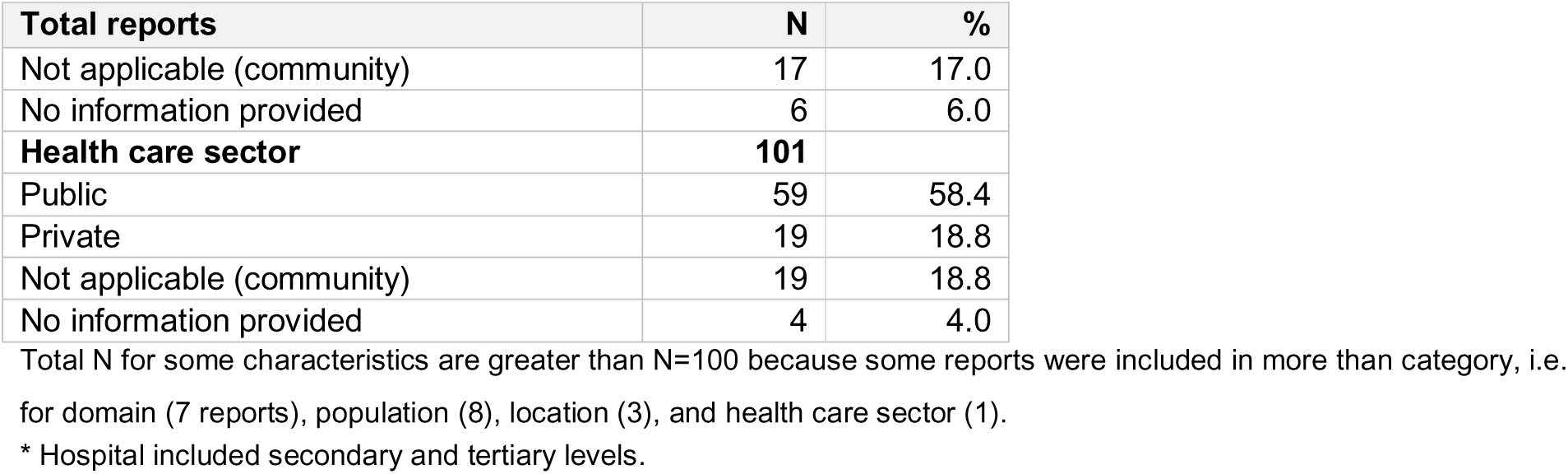
Characteristics of included reports

| Total reports | N | % |
| --- | --- | --- |
|  | **100** |  |
| **Domains** | **107** |  |
| Antibiotic consumption | 20 | 18.7 |
| Appropriateness of antibiotic prescribing | 49 | 45.8 |
| Gyssens method | 18 | 16.8 |
| Reference guidelines | 31 | 29.0 |
| Antibiotic stewardship | 13 | 12.1 |
| Knowledge, attitude and practice survey | 25 | 23.4 |
| **Study design** | **100** |  |
| Observational | 86 | 86.0 |
| Pre-post design | 13 | 13.0 |
| Randomized control trial | 1 | 1.0 |
| **Study population** | **108** |  |
| Patients | 74 | 68.6 |
| Inpatients | 56 | 51.9 |
| Outpatients | 18 | 16.7 |
| Community | 20 | 18.5 |
| Healthcare providers | 14 | 12.9 |
| **Study location** | **103** |  |
| Java | 62 | 60.2 |
| Sumatra | 16 | 15.5 |
| Papua and Nusa Tenggara | 8 | 7.8 |
| Sulawesi | 7 | 6.8 |
| Kalimantan | 6 | 5.8 |
| Bali | 2 | 1.9 |
| National | 2 | 1.9 |
| **Year of study** | **100** |  |
| 2016-2021 | 62 | 62.0 |
| 2011-2015 | 29 | 29.0 |
| 2000-2010 | 9 | 9.0 |
| **Healthcare level** | **100** |  |
| Primary | 24 | 24.0 |
| Hospital* | 53 | 53.0 |
| Not applicable (community) | 17 | 17.0 |
| No information provided | 6 | 6.0 |
| **Health care sector** | **101** |  |
| Public | 59 | 58.4 |
| Private | 19 | 18.8 |
| Not applicable (community) | 19 | 18.8 |
| No information provided | 4 | 4.0 |

Total N for some characteristics are greater than N=100 because some reports were included in more than category, i.e. for domain (7 reports), population (8), location (3), and health care sector (1).
\* Hospital included secondary and tertiary levels.

We included original papers reporting on one or more of the following domains:

1. Antibiotic consumption: estimates of antibiotic utilization in adults expressed as defined daily dose (DDD) normalized to 100 patient-days (inpatients) or inhabitant-days (outpatients) (or with alternative units of measure that could be converted). DDD is defined as the assumed average maintenance drug dose per day for its main indication in adults as established by the WHO Collaborating Centre for Drug Statistics and Methodology. For this review, we extracted data on the top-15 antibiotics with the highest DDDs (defined as the top-3 antibiotics reported in at least two studies) and grouped them according to the WHO AwaRe (Access, Watch, and Reserve) classification.^11^ We excluded paediatric studies because most studies incorrectly used DDD (not suitable for children) rather than days of therapy (DOT).
2. Appropriateness of antibiotic prescribing for treatment or prophylaxis: a) drug prescription audits using Gyssens method,^12, 13^ i.e., an expert panel (of at least two reviewers) retrospectively evaluates each antibiotic prescription (based on available clinical and microbiological data in the medical record and local guidelines) by sequentially categorizing six indicators: (vi) insufficient data; (v) antibiotic is not indicated; (iv) alternative antibiotic is available that is (iv-a) more effective, (iv-b) less toxic, (iv-c) less costly, (iv-d) has narrower spectrum; (iii) inappropriate duration, (iii-a) either too long, (iii-b) or too short; incorrect (ii-a) dose, (ii-b) interval, (ii-c) route; (i) incorrect timing; and (0) appropriate use; b) studies that assessed antibiotic prescriptions according to named reference guideline(s), based on the following eight pre-defined indicators: no contra-indication or allergy label, indication, drug choice, dose, frequency, duration, route of administration, and overall rational use. We included in this category relevant indicators collected as part of point prevalence surveys (e.g., GLOBAL-PPS, WHO).
3. AMS interventions: intervention studies with a clearly described pre-post or (quasi-) experimental design and outcome measures.
4. Knowledge, attitudes and perceptions (KAP): questionnaire-based surveys on knowledge, attitude and/or practice of antibiotic use among consumers and/or providers.

We excluded non-human studies; studies exclusively describing other (non-J01) drugs; describing or comparing the effectiveness, cost, quality and/or molecular profiles of antibiotics; exclusively targeting particular diagnoses and its treatment; and studies of which the full text could not be accessed. Because the included studies were mostly neither randomised controlled studies nor comparative studies, traditional methods for assessment of risk of bias were not applicable. To ensure quality, we excluded studies that did not report an essential set of STROBE checklist core items,^14^ or did not fulfil additional criteria (**Table S2**). All steps in this systematic literature search were conducted using Mendeley reference management software. Two reviewers (PD, MA) separately screened all titles and abstracts, and removed duplicates. Full-text articles were independently judged for relevance and quality by at least two reviewers (PD, MA, MM, RL). Any disagreements were resolved by a senior researcher (RLH).

**Table 2.**
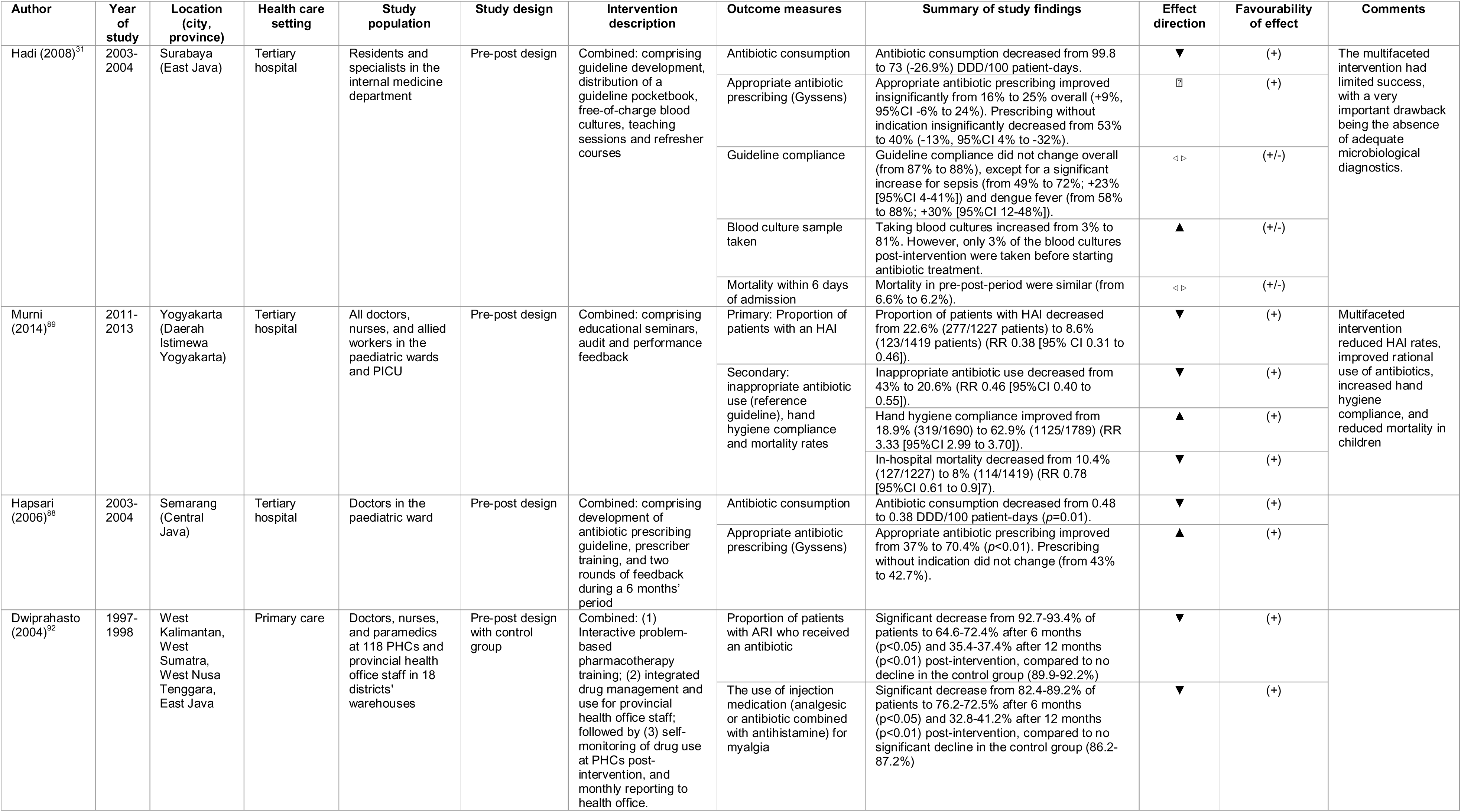
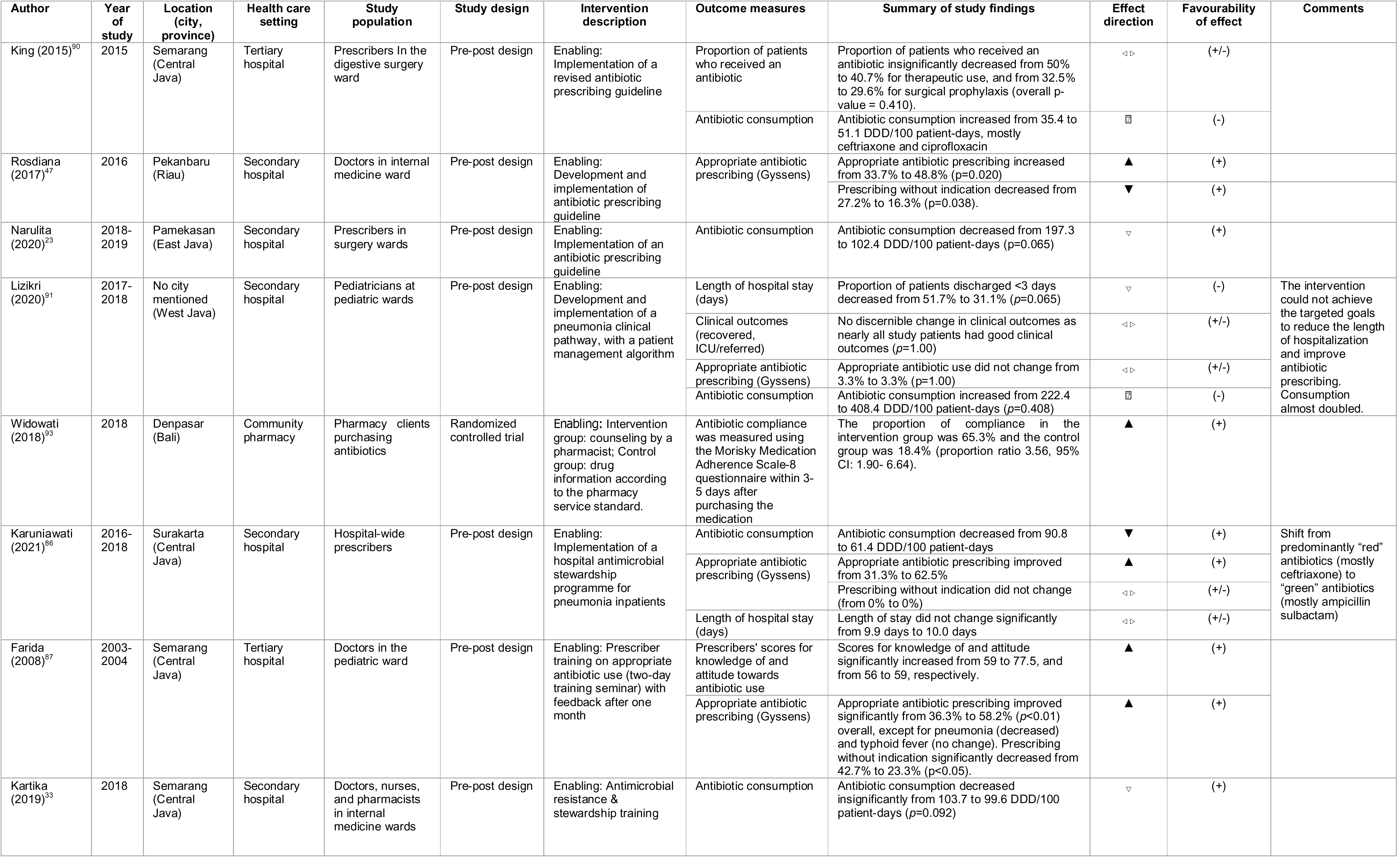
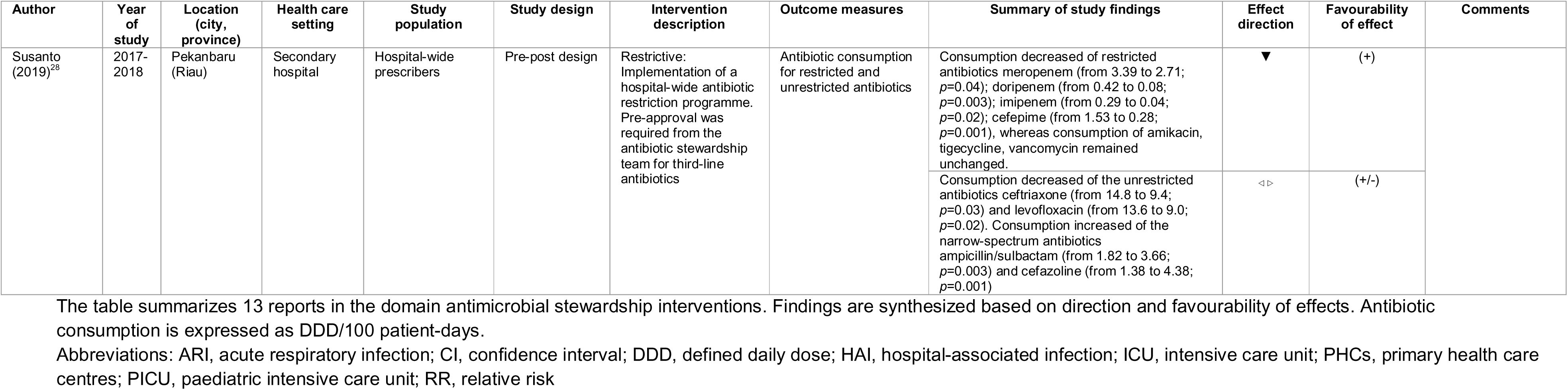
Summary of reports on antimicrobial stewardship interventions

### Data analysis

We extracted and tabulated data on study design, location, population, health care setting, year of study, study sample size, indicators, interventions, outcome measures and effects, and emerging themes, as appropriate and relevant, based on the author-reported summary estimates (not individual patient-level data). Data were extracted by two reviewers (PD, MM) onto a predesigned form.

Where possible, data on antibiotic consumption and appropriateness of antibiotic prescribing were pooled using a random-effects model meta-analysis, due to the likeliness of unexplained heterogeneity.^15^ Data on antibiotic consumption were pooled with meta-analyses of rates using the generic inverse variance method, with DDD/100 patient-days as the rate and sample sizes as the denominator. Data on the appropriateness of antibiotic prescribing, separate for Gyssens method and reference guidelines, were pooled with meta-analyses of proportions using the generalised linear mixed model (GLMM) with the logit transformation to retain studies with extreme proportions.^16^ Heterogeneity was assessed using I^2^ statistic (low <25%, moderate 25-49%, substantial 50-74%, or high 75-100%) and chi-squared test (p<0.10). When two or more studies involved overlapping populations, data synthesis was prioritised to studies with larger sample sizes. Subgroup analyses were then performed by dichotomising the studies based on health care setting (primary care vs hospital [including secondary and tertiary levels]), study location (Java vs non-Java), year of study (before vs after 2016), AWaRe classification (access vs watch antibiotics) (for DDD only), and age groups (adults vs children) (for appropriateness only). To assess the robustness of the findings, we also performed a leave-one-out sensitivity analysis (for DDD and Gyssens), and by replicating the analysis using other transformations (i.e., Freeman-Tukey double arcsine, arcsine, inverse-variance logit, log transformations) and without transformation (for Gyssens only). All meta-analyses were performed in R version 4.1.0 using the additional *meta* (4.18-2)*, metafor* (3.0-2), and *forestplot* packages (1.10.1). When appropriate (k≥10), publication bias assessments were performed visually by using funnel plots and quantitatively by using Egger’s tests with a significance level set at 10%. The funnel plots were generated by plotting the inverse square root of study sizes against the effect estimates due to the expected variability of study sizes and the fact that conventional funnel plots may be inaccurate in assessing potential publication bias in meta-analyses of proportions.^17^

Due to substantial clinical heterogeneity in studies evaluating AMS interventions, we synthesised the findings using a vote-counting method based on direction of effects, and subsequently summarised the results into an effect direction table. The interventions were classified as structural, enabling, persuasive, restrictive, or combined (bundled).^18^ Perception surveys were qualitatively synthesised based on the main emerging themes, stratified by consumers and providers.

### Patient and public involvement statement

The patients and public were not involved in the development of this study.

## RESULTS

### Study characteristics

The search strategy collectively gave 9 323 hits. After title and abstract screening and duplicate removal, 551 articles remained (**Figure 1**). After full-text screening and quality assessment, 100 reports (covering 97 studies) were included. Study characteristics are summarized in **Tables 1** and **S3**. Most studies had an observational design (86.0%, 86/100), were conducted in the public sector (58.4%, 59/101), in hospital settings (53.0%, 53/100), in Java (60.2%, 62/103), and during the most recent five years (62.0%, 62/100).

**Figure 1.**
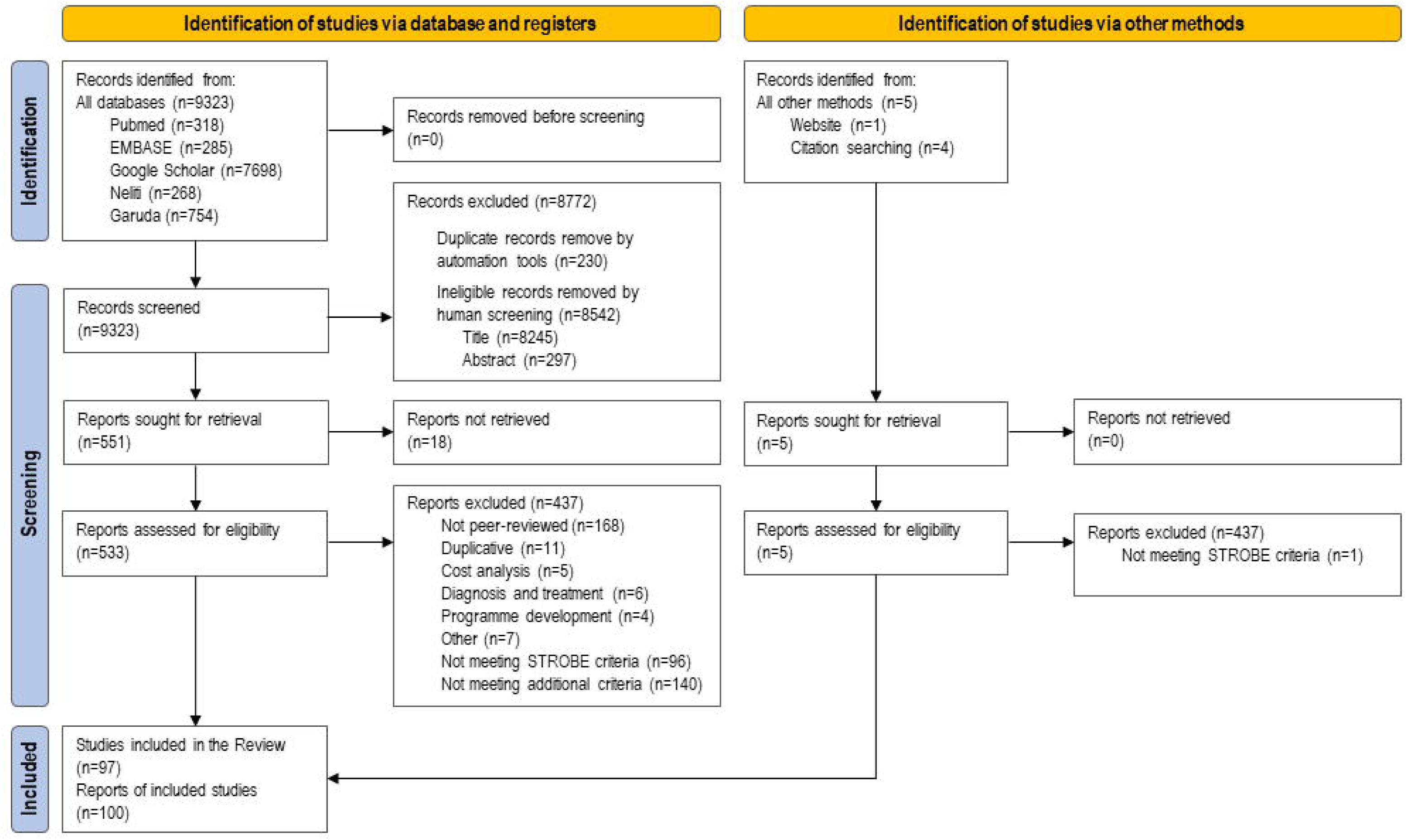
PRISMA flowchart of study selection

**Figure 2.**
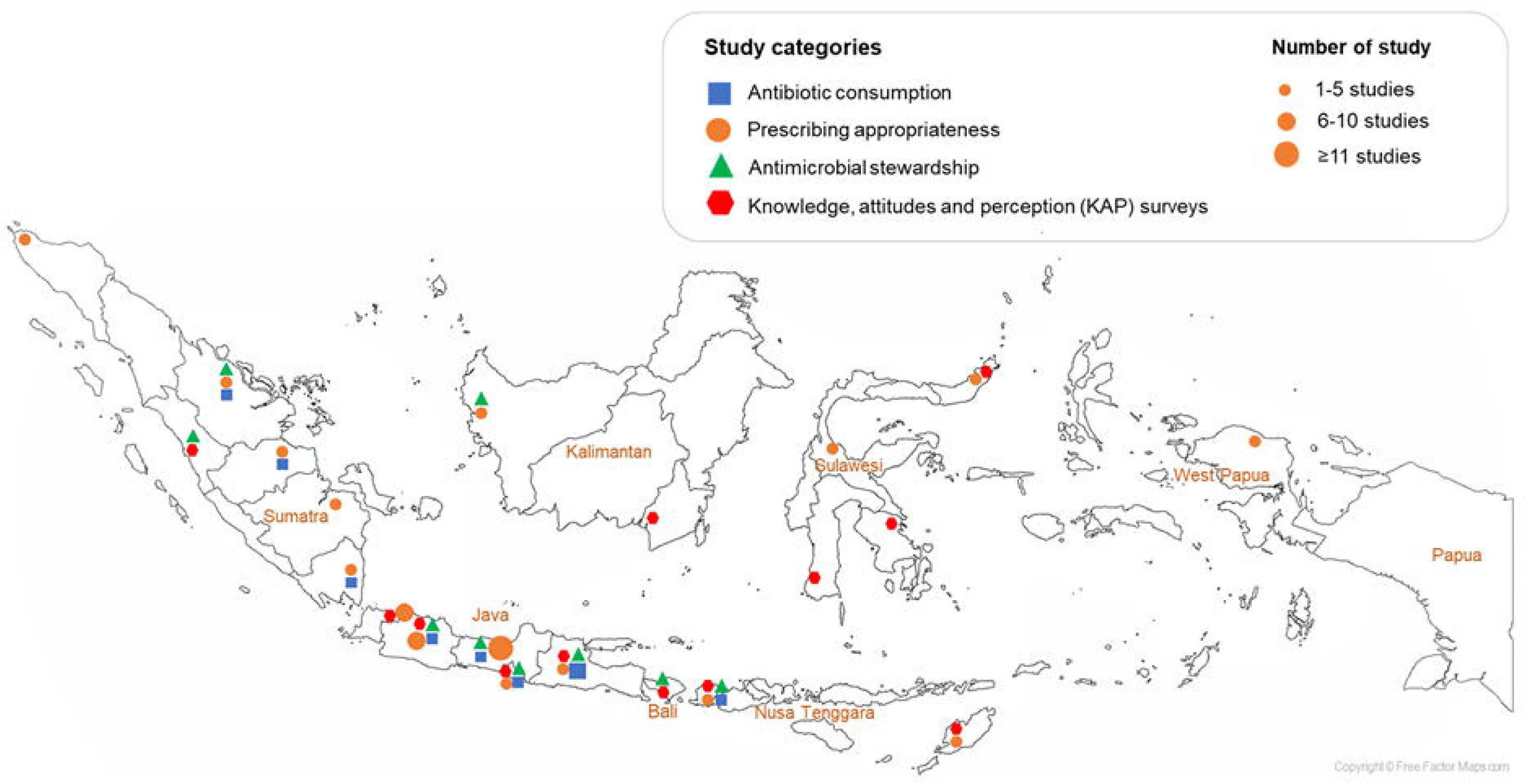
Geographical map of the 100 included reports on antibiotic use in Indonesia 2000-2021 The map includes 2 KAP surveys that were conducted nationwide, and 1 AMS study and 1 KAP survey that were conducted in multiple provinces.

### Antibiotic consumption

A total of 20 reports (5 193 626 patients) reported data on antibiotic consumption.^19–38^ Most of the studies were conducted in hospital settings (16 reports, 76.2%), in Java (14 reports, 66.7%), and between 2016-2021 (17 reports, 81.0%; **Table S4**). The pooled estimate of overall antibiotic consumption was 110.1 DDD/100 patient-days (95% confidence interval [CI] 98.5-121.6; I^2^=100%; **Figure S1**), with ceftriaxone being the most consumed antibiotic (60.2 DDD/100 patient-days [95%CI 34.2-86.1]), followed by ampicillin (28.0 DDD/100 patient-days [95%CI 0.00-82.8]) and levofloxacin (21.4 DDD/100 patient-days [95%CI 15.4-27.3]; **Figure 3**). Sensitivity analysis found that the pooled estimate was not dominated by a single report (**Figure S2**).

**Figure 3.**
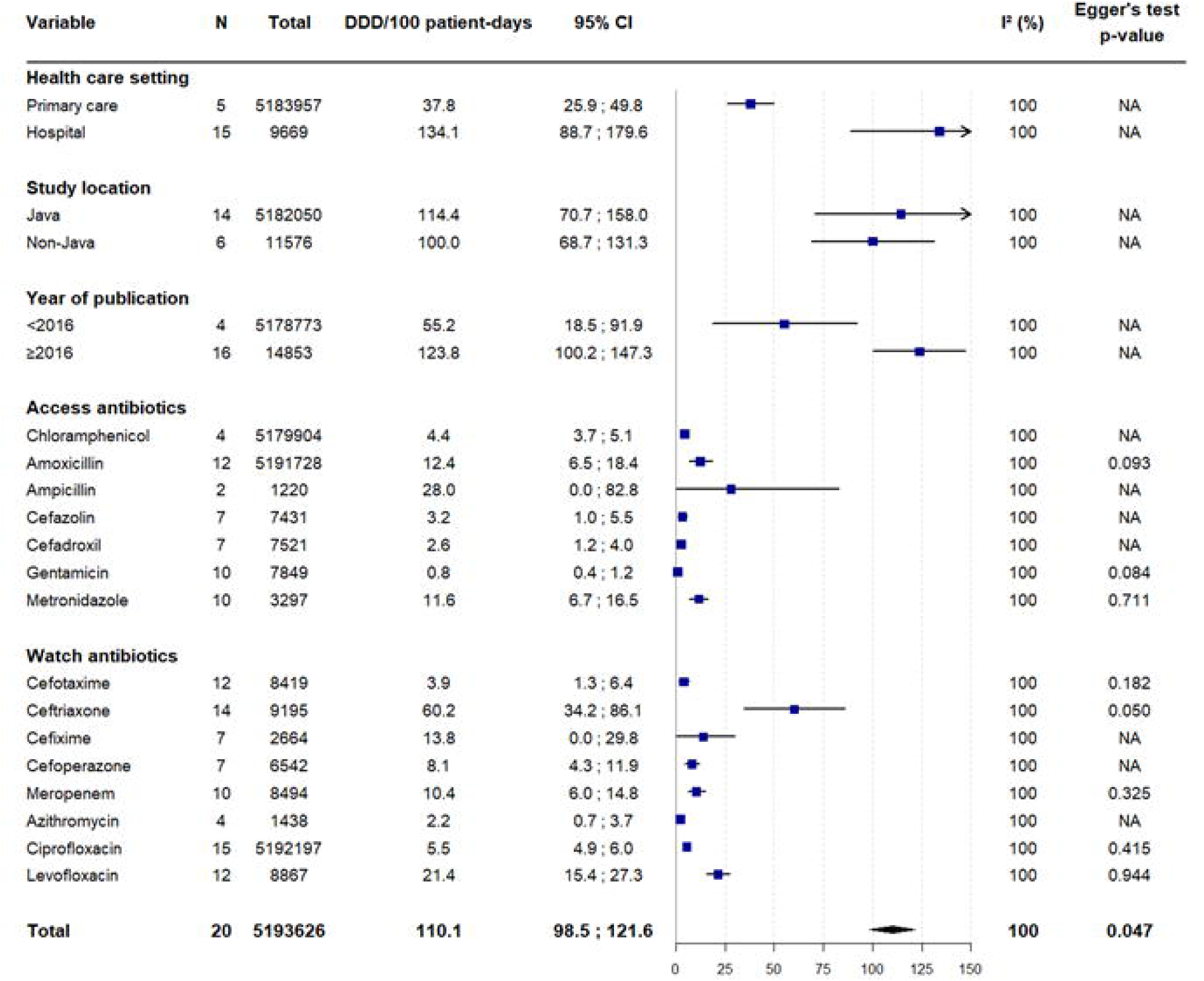
Summary forest plot of reports on antibiotic consumption The figure summarizes 20 reports in the domain antibiotic consumption, expressed as DDD/100 patient-days, for the top-15 antibiotics. Egger’s tests to assess publication bias could only be performed for the primary analysis and the individual AWaRe antibiotics. Abbreviations: CI, confidence interval; DDD, defined daily dose; NA, not applicable.

Subgroup analyses (**Figure 3**) showed higher antibiotic consumption in hospitals (134.1 DDD/100 patient-days [95%CI 88.7-179.6]) than primary care (37.8 DDD/100 patient-days [95%CI 25.9-49.8]), and in recent years (2016-2021: 123.8 DDD/100 patient-days [95%CI 100.2-147.3]) than previous years (2000-2015: 55.2 DDD/100 patient-days [95%CI 18.5-91.9]), but there were no significant differences between Java (114 DDD/100 patient-days [95%CI 70.7-158.0]) and outside of Java (100.0 [95%CI 68.7-131.3]). The asymmetrical funnel plot (Egger’s p=0.047; **Figure S3**) may indicate reporting bias or true heterogeneity in antibiotic consumption patterns between studies, hospitals and/or geographic areas.

### Appropriateness of antibiotic prescribing

A total of 49 reports reported data on the appropriateness of antibiotic prescribing, of which 18^33, 39–55^ (3167 prescriptions) used Gyssens method and 31 (7826 prescriptions)^20, 56–85^ used reference guidelines. Most were conducted in hospitals (37 reports, 74.0%), in Java (33 reports, 66.0%), and in adults (29 reports, 58.0%) (**Table S5-S6**).

Based on the Gyssens method, the pooled estimate for the appropriateness of antibiotic prescribing was 35.3% (95%CI 20.7-53.4%; I^2^=97.2%) (**Figure S4**). confidence analysis showed that the overall estimate was not dominated by a single study (**Figure S5**) and was similar between approximation methods (**Figure S6**). The symmetrical funnel plot (Egger’s p=0.108; **Figure S7**) indicated low risk of bias from small-study effects. Subgroup analyses showed higher appropriateness of antibiotic prescribing in adults (43.1% [95%CI 25.9-62.2%] than in children (8.4% [95%CI 0.6-57.9%]), but there were no significant differences between primary care 49.4% [95% CI 23.7-75.4%]) vs hospitals (32.3% [95%CI 17.9-51.0%]), Java (31.4% [95%CI 18.8-47.6%]) vs outside of Java (38.1% [95%CI 6.2-85.0%], and recent years (2016-2021: 39.1% [95%CI 19.4-63.0%] vs previous years (2000-2015: 26.1% [95%CI 12.4-47.1%] (**Figure 4A**).

**Figure 4.**
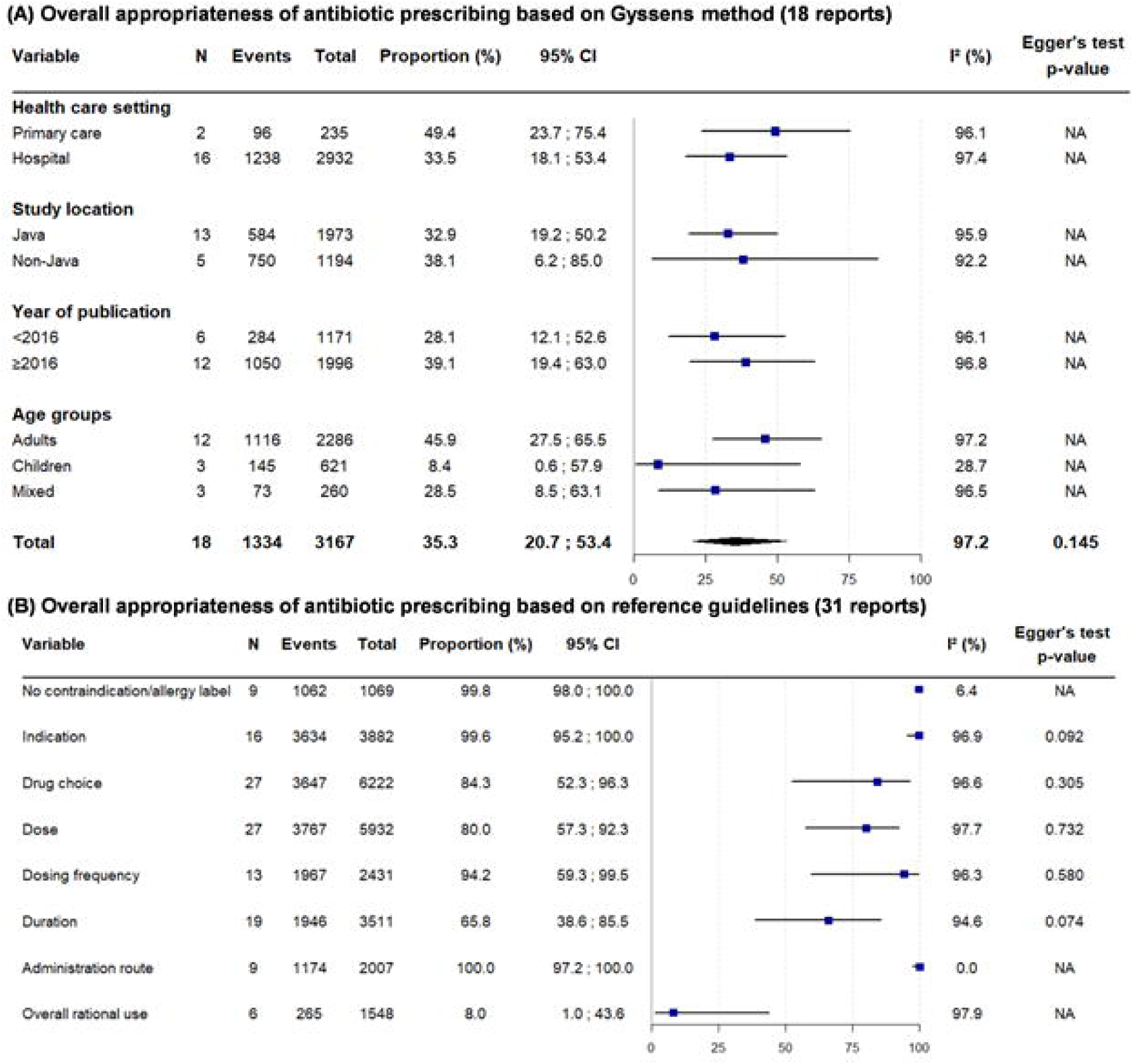
Summary forest plot on prescribing appropriateness The figure summarizes 49 reports in the domain prescribing appropriateness based on Gyssens method (A) or references guidelines (B). Indicators (B) were reported by different numbers of studies. Egger’s tests to assess publication bias in Gyssens method (A) could only be performed for the primary analysis. Abbreviations: CI, confidence interval; NA, not applicable.

Based on reference guidelines, most individual indicators of appropriateness scored excellent (>80%), except for duration (65.8% [95%CI 38.6-85.5%]) (**Figure 4B**) and overall rational antibiotic use (8.0% [95%CI 1.0-43.6%]). Between-study heterogeneity was substantial (75-100%) for all indicators, except for administration route (I^2^=0%, p>0.999) and no contraindication/allergy label (I^2^=6.4%, p=0.382). Possible small-study effects were found for indication and duration (Egger’s test p=0.092 and p=0.074, respectively). Subgroup analysis showed higher appropriateness of duration in hospitals (80.0% [95%CI 66.0-89.2%]) than primary care (34.5% [95%CI 4.3-86.1%]), and in adults (83.2% [95%CI 65.3-92.9%] than children (17.9% [95%CI 1.2-79.4%]; **Figure S8**), and higher appropriateness of drug choice, dosing, and overall rational use outside of Java (95.2% [95%CI 73.9-99.3%], 90.0% [95%CI 71.1-97.1%], and 34.5% [95%CI 3.1-89.8%] than in Java (70.0% [95%CI 23.2-94.7%], 67.2% [95%CI 32.3-89.8%), and 3.3% [95%CI 0.3-32.0%]; **Figure S9-S11,** respectively) –although the latter differences were not statistically significant. There were no significant differences for no contraindication/allergy label, indication, dosing frequency, or administration route (**Figure S12-15**).

### Antibiotic stewardship interventions

A total of 13 reports reported data on the effect of AMS interventions, of which 11 in hospitals,^23, 28, 31, 33, 47, 86–91^ 1 in primary care,^92^ and 1 in community pharmacies^93^ (**Table 2)**. Twelve reports used a pre-post design, and one was a randomized controlled trial. Four reports evaluated a bundled intervention and 9 studies evaluated single interventions. Enabling interventions were most common (antibiotic prescribing guidelines or clinical pathway [8 reports], education and training [5] and pharmacist counselling [1]), followed by persuasive interventions (audit and performance feedback [3]), restrictive interventions (antibiotic restriction with pre-approval [1]) and structural interventions (free blood cultures [1], integrated drug management [1]). The most common reported outcome measures were changes in appropriateness of prescribing (10 reports), antibiotic consumption (8) and mortality (2), followed by prescribers’ scores for knowledge of and attitude towards antibiotic use, blood culture sampling, hand hygiene compliance, clinical outcomes, length of hospital stay, and antibiotic compliance (1 each).

Studies that evaluated bundled interventions (4 reports) reported favourable effects on antibiotic consumption, prescribing appropriateness, guideline compliance, blood culture sampling, HAI rates, hand hygiene compliance, and mortality, although the authors of one study concluded that the multifaceted intervention had limited success, with a very important drawback being the absence of adequate microbiological diagnostics.^31, 88, 89, 92^

Studies that evaluated the implementation of antibiotic prescribing guidelines or clinical pathway (7 reports) reported mixed effects on antibiotic consumption and favourable effects on prescribing appropriateness.^23, 31, 47, 86, 88, 90, 91^ Studies that evaluated education and/or training interventions (5 reports) reported mixed effects on antibiotic consumption and prescribing appropriateness.^31, 87–89, 92^

The one study that evaluated antibiotic restriction with pre-approval found that consumption of restricted antibiotics decreased and of unrestricted narrow-spectrum antibiotics increased.^28^ The one randomised study that evaluated the effect of a pharmacist counselling session of outpatient antibiotic users in community pharmacies found that self-reported antibiotic adherence was significantly higher in the intervention group compared to the control group.^93^

### Knowledge, attitudes, and perceptions on antibiotic use among consumers and providers

A total of 25 reports reported data on KAP, of which 22 among communities^94–115^ and 3 among healthcare providers^116–118^ **(Table S7-S8)**. Interpretation was challenged by the considerable between-study clinical (e.g., study populations) and methodological (e.g., survey questionnaires) heterogeneity.

First, there was a substantial lack of AMR awareness (10 reports) and knowledge about antibiotics (16) among community respondents, with wide variations between communities: overall, 23-26% did not know that antibiotic treated bacterial infections and 58-74% stated that antibiotics can cure viral infections.^96, 105, 110, 115^ Antibiotic knowledge was found to be associated with higher education and higher income (2 reports).^105, 111^

Second, antibiotic self-medication without prescription was reportedly common community respondents, ranging from 20-100% across studies (9 reports). Respondents reported they purchased antibiotics for self-medication at the pharmacy (46-90%),^94–96, 99, 104, 106, 109, 112^ at the kiosk (20-44%),^96, 104^ or received them from family and friends (9-12%).^95, 109^ 20-100% of community respondents reported that they had ever self-medicated with an antibiotic (11),^94– 96, 99, 102, 104, 106, 109, 112–114^ and 87-100% had ever purchased an antibiotic without a prescription (3).^104, 109, 112^ Higher education and higher income was correlated with better antibiotic use knowledge.^111^ One study found that people without health insurance were more likely to self-medicate than those with health insurance,^111^ whereas another study reported the opposite.^104^ The main reasons for self-medication included positive previous experience (54-82%),^102, 104, 109, 112^ self-medication being practical (61-83%),^94^ easy access from the pharmacy (71%),^102^ and doctor visit being expensive (44-72%)^102, 109^ or unpractical (56%).^96^ The main advisors to self-medicate included health care providers (51-83%),^100, 103, 104, 109^ family, relatives or friends (21-45%),^96, 102, 112^ internet (71%),^115^ or reliance on their own knowledge (71%).^96^ Antibiotic adherence levels were not associated with education level or employment status (2 studies).^98, 101^

Third, antibiotic dispensing without prescription was the most important theme reported among health care providers, with conflicting findings. A survey among 250 community pharmacists in Yogyakarta (Java), 68% reported that they would dispense antibiotics without prescription,^116^ whereas a survey among 110 health providers in community health centres in Padang (Sumatera) found that 98.8% did not prescribe antibiotics without a prescription, despite patient request.^117^

## DISCUSSION

This is the first comprehensive, systematic assessment of human antibiotic use in Indonesia, including 100 peer-reviewed reports spanning the past 20 years, covering the key domains of antibiotic consumption, prescribing appropriateness, AMS interventions, and perceptions among consumers and providers. The rising number of scientific reports published in the most recent years in Indonesia reflects the increasing momentum of AMR on the national health agenda. The evidence collected in this Review comes from a range of health care and community settings, including hospitals, primary care, pharmacies and communities, and includes a range of interventions targeting different types of health providers and consumers. Nonetheless, the evidence base is uneven with hospital and urban contexts over-represented, and informal and formal private health providers, who play a major role in antibiotic distribution, particularly underrepresented.

The data included in this Review highlighted several critical evidence gaps. First, there were important limitations in data heterogeneity and study methodology, and publications were predominantly from Java Island, which limited our ability to draw firm conclusions on the contemporary nationwide antibiotic use situation in Indonesia. This is especially relevant given the substantial within-country variations in access to quality health care.^5^ Representative, high-quality data will be essential for benchmarking between provinces, districts, and healthcare facilities, and internationally. Second, there is a lack of knowledge of health system drivers of community antibiotic use, including the effects of the national health insurance rolled-out since 2014, enforcement of antibiotic regulations in informal and private health sectors, as well as how to achieve effective community antibiotic stewardship. Antimicrobials are presently widely available without a prescription, despite existing restrictive policies.^119^ Third, the COVID-19 pandemic has suggested a widespread failure of AMS globally with the potential to worsen the AMR crisis.^4, 120^ Data of its impact in the Indonesian context are urgently needed.

### Antibiotic consumption

Across the included studies, ceftriaxone, levofloxacin (both Watch) and ampicillin (Access) were found to be the most consumed antibiotics among adults, and consumption (expressed as DDD) was higher in hospitals than in primary care, and in recent years (2016-2021) than previous years (2000-2015), but there were no significant differences between geographic settings. According to national pharmaceutical sales data, antibiotic consumption increased 2.5-fold between 2000 and 2015, largely driven by the major classes broad-spectrum penicillins (2.6-fold), fluoroquinolones (7.1-fold), and cephalosporins (5.1-fold).^121^ In 2015, the antibiotic consumption rate per capita in Indonesia (3022 DDDs per 1000 inhabitants per year) fell in the same range as, for instance, China (3060) and Philippines (2600), but was still lower than, for instance, Vietnam (11 480), Thailand (6682) and Malaysia (4388).^121^ About 69% of antibiotic consumption in Indonesia were Access antibiotics, 30% Watch, and <1% Reserve −this is still above the WHO target of 60% Access antibiotics in total consumption.^3^

These trends reflect both better access to antibiotics for those who need them, as well as increases in inappropriate antibiotic use. The findings are consistent with the widespread use of broad-spectrum antibiotics, predominantly third-generation cephalosporins and fluoroquinolones, as reported in other Asian countries^122–126^ and globally,^127, 128^ and with the disproportionate increase in Watch antibiotic consumption observed in LMIC (165% compared with 27.9% in high-income countries between 2000-2015).^129^ Although antibiotic consumption per capita in Indonesia and many other LMIC is currently still lower than in most high-income countries in Europe and North America, rapid increases are leading to converging consumption rates, especially regarding last-resort compounds.^3^ Barring policy changes, antibiotic consumption is projected to increase worldwide by 200% between 2015 and 2030.^3^ The above concerning developments highlight significant challenges for AMS, underscoring the urgent need for regulation of antibiotic use in Indonesia, and other LMIC.

### Appropriateness of prescribing

In Indonesia, appropriateness of antibiotic prescribing was found to be poor overall (35.3%), 49.4% in primary care versus 33.5% in hospitals, 45.9% in adults versus 8.4% in children. We did not identify clear trends over time (prior vs after 2016) or space (Java vs outside of Java). This was despite nationwide implementation of hospital AMS programmes during the past five years.^7^ Studies in primary healthcare settings in other countries have also reported considerable rates of inappropriate antibiotic use, for instance, at 15% in Canada,^130^ 55% in South Africa,^131^ 61% in China,^132^ 88% in Pakistan,^133^ and 90% in Vietnam,^134^ and, likewise, a systematic review of nine studies in LMIC reporting a wide range of between 8 to 100%.^135^ In a global point prevalence survey, guideline compliance of antimicrobial drug choice in hospitals in Latin America, Africa and Asia, was estimated to be lower than 70% for each region.^127^

Inappropriate prescribing of antibiotics has been attributed to a range of complex factors, with variations across settings and countries, including physicians’ nonadherence to antibiotic guidelines, lack of knowledge and training regarding antibiotics, lack of diagnostic facilities or, where available, lack of utilization and quality, uncertainty over the diagnosis or fear of clinical failure, pressure from pharmaceutical industry, financial benefits for physicians, and pressure from patients to prescribe antibiotics regardless of the indication, coupled with lack of time of physicians to educate patients.^136, 137^ Around 60% of total health care spending in Indonesia is in the private sector, where financial incentives may promote prescribing practices that deviate from guidelines. The scale and consequences of non-prescription and private sector antibiotic consumption in Indonesia are an urgent priority for further study and action. Additionally, the local implementation of global definitions and universally applicable quality indicators for antibiotic prescribing in inpatient and outpatient settings will be essential to identify targets for AMS interventions and measure their effectiveness.^138, 139^ Standardized point prevalence survey tools to assess antibiotic use in hospitals have proven to be useful for identifying targets for improvement and evaluating the effect of interventions.^127, 128^

### Antimicrobial stewardship

According to WHO, AMS is considered one of three pillars of an integrated approach to health systems strengthening and reports from high-income settings have shown that AMS can optimise the use of antimicrobials, improve patient outcomes, reduce AMR and health-care-associated infections, and save health-care costs amongst others.^18, 140^ A global systematic review and meta-analysis of 26 studies, reported that implementation of AMS programmes resulted in a reduction in overall antibiotic consumption of 19.1% (US 19.9, Europe 20.9%, Asia 16%), of restricted antibiotics by 26.6%, reduction in drug resistant infections, with no associated adverse outcomes in terms of mortality and infection rates.^141^

In Indonesia, AMS programmes are typically in an early stage of implementation,^142, 143^ with many hospitals lacking the basic infrastructure to adequately measure process, outcome and structural indicators, e.g., access to microbiology services, hospital antibiotic guidelines, AMS staff training and education, human resources (including infectious disease specialist or clinical microbiologist), and ICT support.^144, 145^ Whereas to date government policy has focused on assessing hospital antibiotic use as an AMS outcome indicator (using DDD and Gyssens method),^145^ a recently launched national guideline incorporated additional outcome measures, such as cost-effectiveness, mortality, and AMR rates.^146^

The few AMS intervention studies conducted to date reported clear benefits from implementing bundled interventions combining antibiotic prescribing guidelines, trainings, review and feedback, restriction with pre-approval, among others. These findings demonstrate that AMS interventions are feasible in the local context and that there is considerable potential for reducing antibiotic consumption, particularly of restricted antibiotics, improving prescribing appropriateness, and reducing prescriptions without indication. However, the interventions evaluated to date were mostly single-centre and short-term (<1 year), and data are lacking about the sustained benefits of AMS programmes in the Indonesian context.

Previous evidence has shown that given the varying priorities and contextual issues in LMIC, such as health system processes, patient demands, varying cultures of care, availability of universal access to quality antimicrobials, laboratory infrastructure and surveillance systems, multipronged interventions combining different restrictive and enabling strategies are most likely to be effective.^147, 148^ Indeed, the available Indonesian data confirm that a stand-alone guideline distribution approach, without implementing additional AMS core elements, may not work.^149^ Further enhancement of post-prescription review and feedback efforts holds potential to decrease antibiotic consumption and antibiotic duration.^150^ For pre-prescription approval, local data suggested that the decreased use of last-resort antibiotics might cause a “squeezing the balloon” phenomenon –the increased use of non-restricted antibiotics due to restrictions of the restricted antibiotics.^151^

### Perceptions on antibiotic use

Key themes identified from perception surveys were a substantial lack of AMR awareness and knowledge about antibiotics, particularly among the poor and lower educated, and widespread antibiotic self-medication without prescription among the general population, and over-the-counter non-prescription antibiotic dispensing in community pharmacies but not in community health centres −although representative data are lacking to quantify this problem and its drivers. The available data corroborate findings of a global review of 38 studies from 24 countries in community pharmacies that over-the-counter non-prescription antibiotics comprised 62% of all dispensed antibiotics, most commonly for urinary tract and upper respiratory tract infections.^152^ A review of 19 studies in Southeast Asia reported a prevalence of self-medication with antibiotics ranging from 7.3 to 85.6% (median 42.6%), highest among men, health students and professionals, with the most common illnesses or symptoms being common cold, sore throat, fever, gastrointestinal tract and respiratory diseases.^153^ Across the included studies, the main reasons for self-medication included positive previous experience, easy access from the pharmacy, doctor visit being expensive or unpractical, and the main advisors to self-medicate included health care providers, family, relatives or friends, internet or reliance on their own knowledge, which largely concurred with findings in other Southeast Asian countries.^153^ A mixed-method study in Indonesia found that drug dispensers were driven to sell non-prescription antibiotic due to strong patient demand, informal drug sellers dispensing medicines, competition between different types of drug outlets, drug outlet owners pushing their staff to sell medicines, and weak enforcement of regulations.^119^

### Country progress in the National Action Plan on AMR and suggested priorities

The National Action Plan on AMR has driven efforts to strengthen microbiological laboratory capacity and data information systems to enable national surveillance,^154^ supported by the government and international funding agencies. Indonesia enrolled in the WHO Global Antimicrobial Resistance and Use Surveillance System (GLASS) and submitted first batches of AMR and antibiotic consumption data in 2020.^155^ In 2021, the Ministry of Health launched new guidelines for antibiotic prescribing and stewardship,^146, 156^ which have also adopted the AWaRe classification. Despite this progress, however, the National Action Plan for AMR has not yet generated the required sustainable capacity to contain AMR. Enhanced coordination and financial support at the country, province and district levels are needed, to prevent that the response to AMR remains uneven and insufficient. Interventions to optimise antimicrobial use need to be based on a health systems approach, beyond AMS only, informed by a broad research base. This includes addressing the wider drivers of antibiotic use –such as inequitable burdens of ill health and fractured care cascades. Optimization of antimicrobial use, on the basis of robust surveillance data, should be a priority of the national agenda for universal health coverage. Both universal health coverage and AMR require strong human-centered care with accessible health care facilities, medicines and diagnostics, with a focus on quality and equity.^5, 157, 158^ This requires further investments in health care infrastructure, training of health workers, community participation and increased health literacy. Modifiable factors related to the patient (e.g., awareness, knowledge) and the health system (e.g., strict policies, medicine quality, financial incentives, infrastructure gaps) need to be further identified and addressed when designing context-specific interventions aimed at curtailing inappropriate antibiotic use.

## CONCLUSION

Critical evidence gaps exist on antibiotic use in Indonesia regarding informal and formal private health care providers, geographic areas outside of Java Island, as well as effective AMS models that consider country-specific socio-cultural, economic and political circumstances. Optimisation of antimicrobial use as a means to tackle AMR should be a priority of the national agenda for universal health coverage. In this respect, the COVID-19 pandemic has taught important lessons on the importance of infectious diseases and the critical need to build resilient health systems.

## Data Availability

All data produced in the present work are contained in the manuscript

## Acknowledgements

EXPLAIN study group: Erni J. Nelwan, Ralalicia Limato, Manzilina Mudia, Helio Guterres, Enty Enty, Ifael Y. Mauleti, Maria Mayasari, Iman Firmansyah, May Hizrani, Anis Karuniawati, Prof Taralan Tambunan, Prof Amin Soebandrio, Decy Subekti, Iqbal Elyazar, Mutia Rahardjani, Fitria Wulandari, Prof Reinout van Crevel, Rogier van Doorn, Vu Thi Lan Huong, Nga Do Ti Thuy, Sonia Lewycka, Prof Alex Broom, Raph L. Hamers.

## Author contributions

RLH conceived the idea for the study. RLH and RL obtained the funding. RL, PD and RLH designed the study protocol and data extraction instrument. PD, GL, MM, and MA collected and verified the data, overseen by RL and RLH. RL, GL and RLH performed the analysis and had full access to all study data. RL, GL, MA and RLH drafted the paper, with critical inputs from AK and HRvD. All authors critically revised the manuscript, and all authors gave approval for the final version to be published.

## Data sharing statement

No additional data are available.

