## Supplementary material for "Optimizing antibiotic use in Indonesia: a systematic review and synthesis of current evidence to inform opportunities for intervention"

**Table and Contents**

| Supplementary Material | Page |
|---|---|
| **Methods** | 3 |
| Table S1. Search terms | 3 |
| Table S2. Quality assessment criteria for study inclusion | 3 |
| **Results** | 5 |
| Table S3. Characteristics of included studies | 5 |
| Table S4. Summary of studies on antibiotic consumption based on Defined Daily Dose (DDD) | 11 |
| Figure S1. Forest plot of 20 reports on the defined daily dose (DDD) of total antibiotic use | 13 |
| Figure S2. Results of leave-one-out sensitivity analysis for meta-analysis of the defined daily dose (DDD) of total antibiotic use. | 13 |
| Figure S3. Funnel plot indicating evidence of publication bias (as shown by asymmetry) for meta-analysis of the defined daily dose (DDD) of total antibiotic use | 13 |
| Table S5. Summary of studies on appropriateness of antibiotic prescribing (according to Gyssens method) | 14 |
| Table S6. Summary of studies on appropriateness of antibiotic prescribing (according to reference guidelines) | 16 |
| Figure S4. Forest plot showing results of meta-analysis on the overall appropriateness of antibiotic prescribing according to Gyssens method (18 reports) | 18 |
| Figure S5. Results of leave-one-out sensitivity analysis for meta-analysis on the overall appropriateness of antibiotic prescribing according to Gyssens method | 18 |
| Figure S6. Results of sensitivity analysis for meta-analysis on the overall appropriateness of antibiotic prescribing according to Gyssens method by using alternative methods other than generalized linear mixed model (GLMM) method | 18 |
| Figure S7. Funnel plot for the meta-analysis on the overall appropriateness of antibiotic prescribing according to Gyssens method | 19 |
| Figure S8. Summary forest plot of 19 reports on the appropriateness of antibiotic prescribing according to the “duration” indicator in the reference guidelines | 19 |
| Figure S9. Summary forest plot of 27 reports on the appropriateness of antibiotic prescribing according to the “drug choice” indicator in the reference guidelines | 20 |
| Figure S10. Summary forest plot of 27 reports on the appropriateness of antibiotic prescribing according to the “dose” indicator in the reference guidelines | 20 |
| Figure S11. Summary forest plot of 6 reports on the appropriateness of antibiotic prescribing according to the “overall rational use” indicator in the reference guidelines | 21 |
| Figure S12. Summary forest plot of 9 reports on the appropriateness of antibiotic prescribing according to the “no contraindication/allergy label” indicator in the reference guidelines | 21 |
| Figure S13. Summary forest plot of 16 reports on the appropriateness of antibiotic prescribing according to the “indications” indicator in the reference guidelines | 22 |
| Figure S14. Summary forest plot of 13 reports on the appropriateness of antibiotic prescribing according to the “dosing frequency” indicator in the reference guidelines | 22 |
| Figure S15. Summary forest plot of 9 reports on the appropriateness of antibiotic prescribing according to the “administration route” indicator in the reference guidelines | 23 |
| Table S7. Summary of studies on knowledge, attitudes and perceptions regarding antibiotic use | 24 |
| Table S8. Summary of findings of the knowledge, attitudes and practice surveys | 26 |
| **Reference list of all reports included in the systematic review** | 32 |

#### METHODS

**Table S1.** Search terms

| Database | Keywords |
|---|---|
| PubMed | ("Anti-Bacterial Agents"[Mesh] OR "Antibiotic Prophylaxis"[Mesh]) OR ((“anti-bacterial”[tiab] OR “antibiotic”[tiab] OR “antimicrobial”[tiab]) AND (“use”[tiab] OR “usage”[tiab] OR “prescription*”[tiab] OR “prescribing”[tiab] OR “consumption”[tiab] OR “dispensing”[tiab] OR “stewardship”[tiab])) AND ("Indonesia"[tiab] OR "Indonesia"[MeSH]) |
| EMBASE | (((antibiotic or antimicrobial or antibacterial) and (us* or consum* or prescri* or dispensi* or stewardship) and indonesia) not animal not environment not plant not cancer not chemotherapy not carriage not etiology not susceptib* not molecular not vaccine not probiotic).ti,ab. |
| Google Scholar | indonesia antibiotic\|antimicrobial use\|usage\|prescription\|prescribing\|consumption\|dispensing\|stewardship -animal -environment -plant -cancer -chemotherapy -carriage -etiology -susceptible -susceptibility -molecular -vaccine -probiotic |
| Neliti | (antibiotik) |
| Garba Rujukan Digital (Garuda) | antibiotik (in title) |

**Table S2.** Quality assessment criteria for study inclusion

| No | Items | Antibiotic consumption | Antibiotic appropriateness | Antimicrobial stewardship | Knowledge, attitudes, practices |
|---|---|---|---|---|---|
| **STROBE checklist core items** | | | | | |
| 1 | Presents key elements of study design early in the paper (item 4) | ● | ● | ● | ● |
| 2 | Describe the setting, locations, and relevant dates, including periods of recruitment, exposure, follow-up, and data collection (item 5) | ● | ● | ● | ● |
| 3 | Give the eligibility criteria, and the sources and methods of selection of participants (item 6) | ● | ● | ● | ● |
| 4 | For each variable of interest, give sources of data and details of methods of assessment (measurement) (item 8) | ● | ● | ● | ● |
| 5 | Explain how the study size was arrived at (item 10) | ● | ● | ● | ● |
| 6 | Report numbers of individuals at each stage of study—e.g., numbers potentially eligible, examined for eligibility, confirmed eligible, included in the study, completing follow-up, and analysed (item 13) | ● | ● | ● | ● |
| 7 | Give characteristics of study participants (e.g., demographic, clinical, social) and information | ● | ● | ● | ● |

| No | Items | Antibiotic consumption | Antibiotic appropriateness | Antimicrobial stewardship | Knowledge, attitudes, practices |
|---|---|---|---|---|---|
| | on exposures and potential confounders (item 14) | | | | |
| **Additional theme-specific items** | | | | | |
| 8 | Minimum study sample size | 50 | 50 | | 100 |
| 9 | DDD method correctly applied [a] | ● | | | |
| 10 | Gyssens flowchart method correctly applied (including evaluation by at least 2 independent expert reviewers) [b] | | ● | | |
| 11 | Reference prescribing guidelines specified | | ● | | |
| 12 | Survey questionnaire/questions available in paper | | ● | | ● |
| 13 | Information that is crucial for data interpretation is missing or conflicting | ● | ● | ● | ● |

Abbreviations: DDD, defined daily dose; [a] Definitions and general considerations available from: https://www.whocc.no/ddd/definition_and_general_considera/; [b] Gyssens IC, J Antimicrob Chemother 1992:30:724–7; Van Der Meer JWM, Clin Microbiol Infect 2001;suppl6:12

### RESULTS

**Table S3**. Characteristics of included studies

**Table S3a**. Antibiotic consumption (20 studies)

| No | Author | Year of publication | Year of Study | City, province | Island | Healthcare level | Healthcare sector | Age groups | Study population | No. of population | Study design |
|---|---|---|---|---|---|---|---|---|---|---|---|
| 1 | **Andriani**[1] | 2020 | 2018-2019 | Jambi, Jambi | Sumatra | Primary | Public | Adults | Outpatients | 462 | Cross-sectional |
| 2 | **Dirga**[2,*] | 2021 | 2017 | Lampung, Lampung | Sumatra | Secondary | Public | Adults | Internal medicine inpatients | 164 | Cross-sectional |
| 3 | **Hadi**[3,*] | 2008 | 2003-2004 | Surabaya, East Java | Java | Tertiary | Public | Adults | Fever inpatients (pre-intervention) | 212 | Quasi-experimental |
| 4 | **Herawati**[4] | 2019 | 2016 | Surabaya, East Java | Java | Secondary | Private | Adults | Surgery inpatients | 343 | Cross-sectional |
| 5 | **Kartika** [5,*] | 2019 | 2018 | Semarang, Central Java | Java | Secondary | Public | Adults | Internal medicine inpatients (pre-intervention) | 50 | Quasi-experimental |
| 6 | **Kusuma**[6] | 2016 | 2013 | Banyumas, Central Java | Java | Secondary | Public | Adults | Ob/Gyn inpatients | 247 | Cross-sectional |
| 7 | **Mahmudah**[7] | 2016 | 2013 | Bandung, West Java | Java | Secondary | Private | Adults | Digestive surgery inpatients | 208 | Cross-sectional |
| 8 | **Massey**[8] | 2021 | 2019 | Mataram, West Nusa Tenggara | Nusa Tenggara | Secondary | Public | Adults | Surgery inpatients | 323 | Cross-sectional |
| 9 | **Muliani**[9] | 2021 | 2019 | Surabaya, East Java | Java | Secondary | Private | Adults | Surgery inpatients | 164 | Cross-sectional |
| 10 | **Narulita**[10,*] | 2020 | 2018-2019 | Pamekasan, East Java | Java | Secondary | Public | Adults | Surgery inpatients (pre-intervention) | 200 | Quasi-experimental |
| 11 | **Perdaka**[11] | 2020 | 2017-2018 | Jambi, Jambi | Sumatra | Primary | Public | Adults | Outpatients | 1255 | Cross-sectional |
| 12 | **Pradipta**[12] | 2015 | 2008-2010 | Bandung, West Java | Java | Primary | Public | Adults | Outpatients | 5,178,106 | Cross-sectional |
| 13 | **Pratama**[13] | 2019 | 2016-2017 | Surabaya, East Java | Java | Secondary | Public | Adults | Surgery inpatients | 463 | Cross-sectional |
| 14 | **Putri**[14] | 2021 | 2016-2018 | Yogyakarta, DIY | Java | Secondary | Private | Adults | Adult inpatients with pneumonia | 251 | Cross-sectional |
| 15 | **Rachmawati**[15] | 2020 | 2017 | Pasuruan, East Java | Java | Secondary | Public | Adults | Internal medicine inpatients | 973 | Cross-sectional |
| 16 | **Sholih**[16] | 2019 | 2018 | Karawang, West Java | Java | Primary | Public | Adults | Inpatients | 81 | Cross-sectional |
| 17 | **Susanto**[17] | 2019 | 2017-2018 | Pekanbaru, Riau | Sumatra | Secondary | Private | Adults | Inpatients (pre-intervention) | 5,319 | Quasi-experimental |

| No | Author | Year of publication | Year of Study | City, province | Island | Healthcare level | Healthcare sector | Age groups | Study population | No. of population | Study design |
|---|---|---|---|---|---|---|---|---|---|---|---|
| 18 | **Trisia**[18] | 2020 | 2017-2018 | Jambi, Jambi | Sumatra | Primary | Public | Adults | Outpatients | 4,053 | Cross-sectional |
| 19 | **Wikantiananda**[19] | 2019 | 2016 | (City NA), West Java | Java | Tertiary | Public | Adults | Intensive Care Unit inpatients | 57 | Cross-sectional |
| 20 | **Yulia**[20] | 2017 | 2016 | Surabaya, East Java | Java | Secondary | Private | Adults | Inpatients | 695 | Cross-sectional |

*Studies included in multiple domains.

Abbreviations: NA, Not applicable/available, DIY, Daerah Istimewa Yogyakarta; Ob/Gyn, obstetrics and gynaecology

#### **Table S3b**. Prescribing appropriateness (49 studies)

| No | First author | Year of publication | Year of study | City, Province | Island | Health care level | Healthcare sector | Age groups | Study population | No. of prescription[a] | Study design |
|---|---|---|---|---|---|---|---|---|---|---|---|
| **Gyssens method** | | | | | | | | | | | |
| 1 | **Adani**[21] | 2015 | 2014 | Semarang, Central Java | Java | Primary | Public | Children | Paediatric inpatients | 173 | Cross-sectional |
| 2 | **Aljufri**[22] | 2021 | 2017-2019 | Semarang, Central Java | Java | Tertiary | Public | Adults | Inpatients with pneumonia | 98 | Cross-sectional |
| 3 | **Anggraini**[23] | 2018 | 2014 | Pontianak, West Kalimantan | Kalimantan | Primary | Public | Mixed | Typhoid inpatients | 62 | Cross-sectional |
| 4 | **Hanifah**[24] | 2018 | 2017 | Semarang, Central Java | Java | Secondary | Private | Mixed | Typhoid inpatients | 98 | Cross-sectional |
| 5 | **Hardiana**[25] | 2021 | 2019 | Jakarta, DKI Jakarta | Java | Tertiary | Public | Adults | Inpatients with pneumonia | 88 | Cross-sectional |
| 6 | **Ibrahim**[26] | 2020 | 2019 | Surabaya, East Java | Java | Tertiary | Public | Adults | Inpatients with infection | 84 | Cross-sectional |
| 7 | **Inez**[27] | 2019 | 2018 | Tanjungpura, West Kalimantan | Kalimantan | Secondary | Public | Children | Inpatients | 63 | Quasi-experimental |
| 8 | **Kartika**[5,*] | 2019 | 2018 | Semarang, Central Java | Java | Secondary | Public | Adults | Internal medicine inpatients | 50 | Quasi-experimental |
| 9 | **Maakh**[28] | 2019 | 2018 | Atambua, East Nusa Tenggara | Nusa Tenggara | Secondary | Public | Adults | Ob/Gyn inpatients | 100 | Cross-sectional |
| 10 | **Magdalena**[29] | 2018 | 2017 | Pekanbaru, Riau | Sumatra | Secondary | Private | Adults | ICU and medical ward inpatients | 877 | Cross-sectional |
| 11 | **Muthoharoh**[30] | 2018 | 2017 | Pekalongan, Central Java | Java | Secondary | Public | Mixed | Surgical inpatients | 100 | Cross-sectional |
| 12 | **Purwaningsih**[31] | 2015 | 2014-2015 | Semarang, Central Java | Java | Secondary | Private | Children | Inpatients | 385 | Cross-sectional |
| 13 | **Rosdiana**[32,*] | 2018 | 2016 | Pekanbaru, Riau | Sumatra | Secondary | Public | Adults | Inpatients | 92 | Quasi-experimental |
| 14 | **Setiawan**[33] | 2018 | 2016 | Surabaya, East Java | Java | Tertiary | Public | Adults | ICU inpatients | 110 | Cross-sectional |

| No | First author | Year of publication | Year of study | City, Province | Island | Health care level | Healthcare sector | Age groups | Study population | No. of prescription[a] | Study design |
|---|---|---|---|---|---|---|---|---|---|---|---|
| 15 | **Sumiwi**[34] | 2014 | 2013 | Bandung, West Java | Java | NA | NA | Adults | Digestive surgery inpatients | 344 | Cross-sectional |
| 16 | **Sutrisno**[35] | 2013 | 2011-2012 | Yogyakarta, DIY DIY | Java | Tertiary | Public | Adults | Pneumonia inpatients | 57 | Cross-sectional |
| 17 | **Waridiarto**[36] | 2015 | 2015 | Semarang, Central Java | Java | Tertiary | Public | Adults | Orthopaedic inpatients | 150 | Cross-sectional |
| 18 | **Yoanitha**[37] | 2018 | 2016 | Bandung, West Java | Java | Tertiary | Public | Adults | Ob/Gyn inpatients | 236 | Cross-sectional |
| **Reference guidelines** | | | | | | | | | | | |
| 19 | **Andrajati**[38] | 2016 | 2012 | Depok, West Java | Java | Primary | Public | Mixed | Outpatients with various indications | 392 | Cross-sectional |
| 20 | **Anggraini**[39] | 2020 | 2013 | Jakarta, DKI Jakarta | Java | NA | Public | Adults | Pre-surgical inpatients | 837 | Cross-sectional |
| 21 | **Bakhtiar**[40] | 2019 | 2018 | Sorong, West Papua | Papua | Secondary | Public | Adults | Post-surgical inpatients | 83 | Cross-sectional |
| 22 | **Benua**[41] | 2019 | 2018 | Poso, Central Sulawesi | Sulawesi | Primary | Public | Mixed | Acute respiratory infection outpatients | 126 | Cross-sectional |
| 23 | **Bestari**[42] | 2017 | 2016 | (City NA), Central Java | Java | Tertiary | Public | Children | Paediatric pneumonia inpatients | 90 | Cross-sectional |
| 24 | **Dania**[43] | 2016 | 2014 | Yogyakarta, DIY | Java | Secondary | Private | Adults | Ob/Gyn inpatients receiving prophylaxis and therapeutic antibiotics | 59 | Cross-sectional |
| 25 | **Dewi**[44] | 2018 | 2017 | Jakarta, DKI Jakarta | Java | Tertiary | Public | Adults | ICU inpatients with sepsis or septic shock | 60 | Cross-sectional |
| 26 | **Dewi**[45] | 2020 | 2018 | Jambi, Jambi | Sumatra | Primary | Public | Children | Acute respiratory infection outpatients | 51 | Cross-sectional |
| 27 | **Dewi**[46] | 2020 | 2019 | Jambi, Jambi | Sumatra | Primary | Public | Children | Acute respiratory infection outpatients | 70 | Cross-sectional |
| 28 | **Dirga**[2,*] | 2021 | 2017 | Lampung, Lampung | Sumatra | Secondary | Public | Adults | Internal medicine inpatients | 168 | Cross-sectional |
| 29 | **Elvina**[47] | 2017 | 2016 | Jakarta, DKI Jakarta | Java | NA | NA | Adults | Inpatients with pneumonia | 96 | Cross-sectional |
| 30 | **Fakhrunnisa**[48] | 2020 | 2018 | Tegal, Central Java | Java | Primary | Public | Mixed | Acute respiratory infection outpatients | 632 | Cross-sectional |
| 31 | **Fithria**[49] | 2015 | 2013 | Semarang, Central Java | Java | Secondary | Private | Children | Inpatients with acute diarrhoea | 54 | Cross-sectional |
| 32 | **Grassella**[50] | 2019 | 2018 | Pontianak, West Kalimantan | Kalimantan | Secondary | Public | Children | Acute respiratory infection outpatients | 340 | Cross-sectional |
| 33 | **Harartasyahrani**[51] | 2021 | 2020 | Prabumulih, South Sumatra | Sumatra | Secondary | Private | Adults | Surgery inpatients | 119 | Cross-sectional |
| 34 | **Hasyul**[52] | 2019 | 2017 | Garut, West Java | Java | Primary | Public | Mixed | Typhoid outpatients | 705 | Cross-sectional |
| 35 | **Herlina**[53] | 2021 | 2017 | Mataram, West Nusa Tenggara | Nusa Tenggara | Secondary | Public | Adults | Urinary tract infection inpatients | 105 | Cross-sectional |
| 36 | **Islam**[54] | 2017 | 2014 | Jakarta, DKI Jakarta | Java | Secondary | Public | Adults | Pneumonia inpatients | 139 | Cross-sectional |
| 37 | **Jamiati**[55] | 2019 | 2017 | Gayo Lues, Aceh | Sumatra | Primary | Public | Mixed | Outpatients | 86 | Cross-sectional |

| No | First author | Year of publication | Year of study | City, Province | Island | Health care level | Healthcare sector | Age groups | Study population | No. of prescription[a] | Study design |
|---|---|---|---|---|---|---|---|---|---|---|---|
| 38 | **Kaparang**[56] | 2014 | 2013 | Manado, North Sulawesi | Sulawesi | Tertiary | Public | Children | Pneumonia inpatients | 112 | Cross-sectional |
| 39 | **Kurniawati**[57] | 2021 | 2019 | Yogyakarta, DIY | Java | Secondary | NA | Adults | Urinary tract infection inpatients | 61 | Cross-sectional |
| 40 | **Limato**[58] | 2021 | 2019 | Jakarta, DKI Jakarta | Java | Secondary, Tertiary | Public, Private | Mixed | Inpatients receiving systemic antimicrobials | 915 | Cross-sectional |
| 41 | **Nawakasari**[59] | 2019 | 2017 | Klaten, Central Java | Java | Tertiary | Public | Adults | Urinary tract infection inpatients | 76 | Cross-sectional |
| 42 | **Nyoman**[60] | 2017 | 2015 | Bandung, West Java | Java | Primary | Private | Children | Acute respiratory infection outpatients | 425 | Cross-sectional |
| 43 | **Octavia**[61] | 2019 | 2018 | Lamongan, East Java | Java | Secondary | Private | Adults | Ob/Gyn inpatients (prophylaxis) | 54 | Cross-sectional |
| 44 | **Ofisya**[62] | 2020 | 2019 | Pontianak, West Kalimantan | Kalimantan | Secondary | Public | Mixed | Pneumonia inpatients | 86 | Cross-sectional |
| 45 | **Oktaviani**[63] | 2015 | 2014-2015 | (City NA), Riau | Sumatra | Secondary | Public | Adults | Ob/Gyn inpatients (prophylaxis) | 140 | Cross-sectional |
| 46 | **Ovikariani**[64] | 2019 | 2019 | Semarang, Central Java | Java | Primary | Public | Mixed | Pneumonia outpatients | 79 | Cross-sectional |
| 47 | **Rusdiana**[65] | 2016 | 2014 | Tangerang, West Java | Java | Secondary | Private | Adults | Ob/Gyn inpatients (prophylaxis) | 256 | Cross-sectional |
| 48 | **Sugiarti**[66] | 2015 | 2014 | Malang, East Java | Java | Primary | Public | Children | Acute respiratory infection outpatients | 120 | Cross-sectional |
| 49 | **Zazuli**[67] | 2015 | 2009 | Bandung, West Java | Java | Secondary | Private | Adults | Pre- and post-surgical inpatients | 1290 | Cross-sectional |

*Studies included in multiple domains. [a] For reference guidelines, we reported as total number of prescriptions the highest number reported in a given domain; if no number of prescriptions was reported, we reported total number of patients.

Abbreviations: NA, Not Applicable/available; DIY, Daerah Istimewa Yogyakarta; DKI, Daerah Khusus Ibukota; Ob/Gyn, Obstetrics and gynaecology

**Table S3c**. Antimicrobial stewardship (13 studies)

| No | First author | Year of publication | Year of Study | City, province | Island | Healthcare level | Healthcare sector | Age groups | Study population | No. of population | Study design |
|---|---|---|---|---|---|---|---|---|---|---|---|
| 1 | **Dwiprahasto**[68] | 2004 | 1997-1998 | West Kalimantan, West Sumatra, West Nusa Tenggara | Kalimantan, Sumatra, Nusa Tenggara | Primary | Public | NA | Doctors, nurses, and paramedics | NA | Quasi-experimental |
| 2 | **Farida**[69] | 2008 | 2003-2004 | Semarang, Central Java | Java | Tertiary | Public | Children | Doctors | 22 | Quasi-experimental |
| 3 | **Hadi**[3,*] | 2008 | 2003-2004 | Surabaya, East Java | Java | Tertiary | Public | Adults | Doctors | 155 | Quasi-experimental |
| 4 | **Hapsari**[70] | 2006 | 2003-2004 | Semarang, Central Java | Java | Tertiary | Public | Children | Doctors | NA | Quasi-experimental |
| 5 | **Kartika**[5,*] | 2019 | 2018 | Semarang, Central Java | Java | Secondary | Public | Adults | Doctors, nurses, and pharmacists | NA | Quasi-experimental |

| No | First author | Year of publication | Year of Study | City, province | Island | Healthcare level | Healthcare sector | Age groups | Study population | No. of population | Study design |
|---|---|---|---|---|---|---|---|---|---|---|---|
| 6 | **Karuniawati**[71] | 2021 | 2016-2018 | Surakarta, Central Java | Java | Secondary | Public | Adults | Doctors | NA | Quasi-experimental |
| 7 | **King**[72] | 2015 | 2014-2015 | Semarang, Central Java | Java | Tertiary | Public | Adults | Doctors | NA | Quasi-experimental |
| 8 | **Lizikri**[73] | 2020 | 2017 | (City NA), West Java | Java | NA | Private | Children | Doctors | NA | Quasi-experimental |
| 9 | **Murni**[74] | 2014 | 2011-2013 | Yogyakarta, DIY | Java | Tertiary | Public | Children | Doctors, nurses, and allied health workers | NA | Quasi-experimental |
| 10 | **Narulita**[10,*] | 2020 | 2018-2019 | Pamekasan, East Java | Java | Secondary | Public | Adults | Doctors | 200 | Quasi-experimental |
| 11 | **Rosdiana**[32,*] | 2018 | 2016 | Pekanbaru, Riau | Sumatra | Secondary | Public | Adults | Doctors | NA | Quasi-experimental |
| 12 | **Susanto**[17] | 2019 | 2017-2018 | Pekanbaru, Riau | Sumatra | Secondary | Private | Adults | Doctors | NA | Quasi-experimental |
| 13 | **Widowati**[75] | 2018 | 2018 | Denpasar, Bali | Bali | Primary, community pharmacy | NA | Adults | Pharmacy visitors | 98 | Experimental (Randomized Control Trial) |

*Studies included in multiple domains
Abbreviation: NA, Not Applicable/available; DIY, Daerah Istimewa Yogyakarta

**Table S3d**. Knowledge, attitudes and perception surveys (25 studies)

| No | First author | Year of publication | Year of Study | City, province | Island | Healthcare level | Healthcare sector | Study population | No. of population | Study design |
|---|---|---|---|---|---|---|---|---|---|---|
| 1 | **Artini**[76] | 2016 | 2014-2015 | Denpasar, Bali | Bali | NA | NA | Community respondents: medical and non-medical university students | 240 | Cross-sectional |
| 2 | **Asvinigita**[77] | 2019 | 2019 | Yogyakarta, DIY | Java | NA | NA | Healthcare providers: community pharmacists | 250 | Cross-sectional |
| 3 | **Djawaria**[78] | 2018 | 2014-2015 | Surabaya, East Java | Java | NA | NA | Community respondents at pharmacy | 267 | Cross-sectional |
| 4 | **Fatmah**[79] | 2019 | 2018 | Mataram, West Nusa Tenggara | Nusa Tenggara | NA | NA | Community respondents: medical and non-medical university students | 400 | Cross-sectional |
| 5 | **Fernandez**[80] | 2013 | 2012 | Manggarai Barat, East Nusa Tenggara | Nusa Tenggara | NA | NA | Community respondents at pharmacies | 108 | Cross-sectional |
| 6 | **Fimanggara**[81] | 2016 | 2013 | Jatinangor, West Java | Java | NA | NA | Community respondents: non-medical college students | 250 | Cross-sectional |
| 7 | **Fitriah**[82] | 2021 | 2019 | Banjarbaru, South Kalimantan | Kalimantan | NA | NA | Community respondents: household members | 380 | Cross-sectional |
| 8 | **Hamid**[83] | 2020 | 2019 | Pangkajene and Kepulauan, South Sulawesi | Sulawesi | NA | NA | Community respondents: teachers in public schools | 236 | Cross-sectional |

| No | First author | Year of publication | Year of Study | City, province | Island | Healthcare level | Healthcare sector | Study population | No. of population | Study design |
|---|---|---|---|---|---|---|---|---|---|---|
| 9 | **Insany**[84] | 2015 | 2014 | Bandung, West Java | Java | Primary | Public | Outpatients from primary health centres and pharmacies | 508 | Cross-sectional |
| 10 | **Kondoj**[85] | 2020 | 2019-2020 | Manado, North Sulawesi | Sulawesi | NA | NA | Community respondents | 290 | Cross-sectional |
| 11 | **Kristina**[86] | 2020 | 2018 | Yogyakarta, DIY | Java | Primary | NA | Outpatients at clinics and pharmacies | 268 | Cross-sectional |
| 12 | **Kurniawan**[87] | 2017 | 2015 | Manado, North Sulawesi | Sulawesi | Primary | Public | Community respondents at primary health centres | 400 | Cross-sectional |
| 13 | **Novelni**[88] | 2020 | 2019 | Padang, West Sumatra | Sumatra | NA | NA | Community respondents | 100 | Cross-sectional |
| 14 | **Nuraini**[89] | 2018 | 2017-2018 | Bangkalan, East Java | Java | Secondary | Public | Outpatients at secondary hospital | 103 | Cross-sectional |
| 15 | **Pratama**[90] | 2018 | 2016 | Jember, East Java | Java | NA | NA | Community respondents: non-medical university students | 324 | Cross-sectional |
| 16 | **Salsabila**[91] | 2020 | 2019 | Yogyakarta, DIY | Java | NA | NA | Community respondents in rural and urban areas | 125 | Cross-sectional |
| 17 | **Siahaan**[92] | 2017 | 2015 | No city mentioned, West Java, DKI Jakarta, Southeast Sulawesi | Java, Sulawesi | NA | NA | Community respondents: household members | 1271 | Cross-sectional |
| 18 | **Siswati**[93] | 2009 | 2000 | Padang, West Sumatra | Sumatra | Primary | Public | Healthcare providers | 110 | Cross-sectional |
| 19 | **Tandjung**[94] | 2021 | 2020-2021 | Manado, North Sulawesi | Sulawesi | NA | NA | Community respondents | 323 | Cross-sectional |
| 20 | **WHO**[95] | 2015 | 2015 | National | National | NA | NA | Community respondents: household members | 1027 | Cross-sectional |
| 21 | **Widayati**[96] | 2011 | 2010 | Yogyakarta, DIY | Java | NA | NA | Community respondents: household members | 559 | Cross-sectional |
| 22 | **Widayati**[97] | 2012 | 2010 | Yogyakarta, DIY | Java | NA | NA | Community respondents: household members | 559 | Cross-sectional |
| 23 | **Yulia**[98] | 2019 | 2019 | Bukittinggi, West Sumatra | Sumatra | Primary | Public | Community respondents | 100 | Cross-sectional |
| 24 | **Yuliani**[99] | 2014 | 2014 | Kupang, East Nusa Tenggara | Nusa Tenggara | NA | NA | Community respondents | 100 | Cross-sectional |
| 25 | **Zhang**[100] | 2020 | 2018 | National | National | NA | NA | Healthcare providers: General practitioners and paediatricians | 100 | Cross-sectional |

Abbreviations: NA, Not Applicable/available; DIY, Daerah Istimewa Yogyakarta; DKI, Daerah Khusus Ibukota

**Table S4.** Summary of studies on antibiotic consumption based on Defined Daily Dose (DDD)

| First author | Year of publication | Location | Health care level | Study population | No. of patients | Total antibiotics | Access Antibiotics | | | | | | | Watch Antibiotics | | | | | | | |
|---|---|---|---|---|---|---|---|---|---|---|---|---|---|---|---|---|---|---|---|---|---|
| | | | | | | | Chloramphenicol | Amoxicillin | Ampicillin | Cefazolin | Cefadroxil | Gentamicin | Metronidazole | Cefotaxime | Ceftriaxone | Cefixime | Cefoperazone | Meropenem | Azithromycin | Ciprofloxacin | Levofloxacin |
| **Secondary/tertiary care** | | | | | | | | | | | | | | | | | | | | | |
| **Dirga**[2] | 2021 | Lampung | Secondary | Internal medicine inpatients | 164 | 116.3 | - | - | - | - | - | 0.5 | 22.4 | 3.9 | 62.3 | 1.8 | 1.5 | 0.4 | 3.9 | 1.5 | 11.7 |
| **Hadi**[3] | 2008 | Surabaya | Tertiary | Fever inpatients (pre-intervention) | 212 | 99.8 | - | - | - | - | - | - | - | - | - | - | - | - | - | - | - |
| **Herawati**[4] | 2019 | Surabaya | Tertiary | Surgery inpatients | 343 | 30.4 | - | - | - | - | - | - | - | - | - | - | - | - | - | - | - |
| **Kartika**[5] | 2019 | Semarang | Secondary | Internal medicine inpatients (pre-intervention) | 50 | 103.7 | - | 0.0 | - | - | - | 0.7 | 0.0 | 3.8 | 45.9 | 1.9 | - | - | 3.2 | 20.8 | 27.4 |
| **Kusuma**[6] | 2016 | Banyumas | Secondary / Tertiary | Ob/Gyn inpatients | 247 | 77.7 | - | 22.7 | 56.0 | 1.0 | 10.7 | - | - | 6.0 | 2.3 | - | 6.0 | - | - | - | - |
| **Mahmudah**[7] | 2016 | Bandung | Tertiary | Digestive surgery inpatients | 208 | 18.0 | - | - | - | 0.0 | 0.1 | 0.0 | 4.6 | 0.3 | 8.8 | 1.1 | 0.1 | 0.4 | - | 0.7 | 0.1 |
| **Massey**[8] | 2021 | Mataram | Secondary | Surgery inpatients | 323 | 414.7 | - | 9.0 | - | - | 2.0 | - | 3.1 | 23.7 | 258.6 | 45.5 | 2.1 | - | - | 2.0 | 16.1 |
| **Muliani**[9] | 2021 | Surabaya | Secondary | Surgery inpatients | 164 | 144.3 | - | 0.7 | - | 5.1 | - | 0.2 | 12.2 | 0.1 | 75.6 | - | - | 20.9 | - | - | 0.8 |
| **Narulita**[10] | 2020 | Pamekasan | Secondary | Surgery inpatients (pre-intervention) | 200 | 197.4 | - | - | - | - | - | 1.7 | 12.7 | 0.1 | 160.2 | - | 0.2 | 1.1 | - | 17.3 | 0.1 |
| **Pratama**[13] | 2019 | Surabaya | Secondary | Surgery inpatients | 463 | 102.9 | - | - | - | 11.3 | - | 1.9 | 27.3 | 2.3 | 53.9 | - | - | 3.3 | - | - | 1.1 |

| First author | Year of publication | Location | Health care level | Study population | No. of patients | Total antibiotics | Access Antibiotics | | | | | | | Watch Antibiotics | | | | | | | |
|---|---|---|---|---|---|---|---|---|---|---|---|---|---|---|---|---|---|---|---|---|---|
| | | | | | | | Chloramphenicol | Amoxicillin | Ampicillin | Cefazolin | Cefadroxil | Gentamicin | Metronidazole | Cefotaxime | Ceftriaxone | Cefixime | Cefoperazone | Meropenem | Azithromycin | Ciprofloxacin | Levofloxacin |
| **Putri**[14] | 2021 | Yogyakarta | Secondary | Inpatients with pneumonia | 251 | 43.8 | - | - | - | - | - | 0.4 | - | 0.1 | 19.5 | 2.8 | - | 0.3 | 1.3 | 0.3 | 4.0 |
| **Rachmawati**[15] | 2020 | Pasuruan | Secondary | Internal medicine inpatients | 973 | 75.3 | - | 0.3 | 0.1 | 0.0 | 4.2 | 0.1 | 9.7 | 2.3 | 27.8 | 5.3 | - | 2.6 | 0.3 | 8.4 | 3.6 |
| **Susanto**[17] | 2019 | Pekanbaru | Secondary | Inpatients (pre-intervention) | 5319 | 49.5 | - | 0.8 | - | 1.4 | - | 0.0 | - | 0.8 | 14.8 | - | 1.0 | 3.4 | - | 0.6 | 13.6 |
| **Wikantiananda**[19] | 2019 | West Java province | Tertiary | ICU inpatients | 57 | 296.8 | - | - | - | 3.8 | - | 2.6 | 17.0 | 2.9 | 30.6 | - | - | 49.9 | - | 3.4 | 143.2 |
| **Yulia**[20] | 2017 | Surabaya | Secondary | Hospital-wide inpatients | 695 | 241.3 | - | 20.6 | - | - | - | - | 7.1 | - | 79.2 | 38.1 | - | 21.9 | - | 9.0 | 34.5 |
| **Primary health care** | | | | | | | | | | | | | | | | | | | | | |
| **Andriani**[1] | 2020 | Jambi | Primary | Outpatients | 462 | 6.9 | 0.0 | 4.0 | - | - | 0.6 | - | - | - | - | - | - | - | - | 1.9 | - |
| **Perdaka**[11] | 2020 | Jambi | Primary | Outpatients | 1,255 | 6.4 | 0.1 | 4.6 | - | - | 0.1 | - | - | - | - | - | - | - | - | 1.5 | - |
| **Pradipta**[12] | 2015 | Bandung | Primary | Outpatients | 5,178,106 | 25.3 | 0.4 | 16.8 | - | - | - | - | - | - | - | - | - | - | - | 1.6 | - |
| **Sholih**[16] | 2019 | Karawang | Primary | Inpatients | 81 | 144.5 | 17.1 | 65.8 | - | - | - | - | - | - | 2.7 | - | 45.9 | - | - | 12.3 | - |
| **Trisia**[18] | 2020 | Jambi | Primary | Outpatients | 4,053 | 6.1 | - | 3.9 | - | - | 0.6 | - | - | - | - | - | - | - | - | 1.1 | - |

Abbreviations: -, not available; ICU, intensive care unit; Ob/Gyn, Obstetrics and gynaecology

The table summarizes 20 studies on antibiotic consumption, expressed as (or converted to) Defined Daily Dose (DDD) per 100 patient-days, overall and/or for individual antibiotics, and divided by primary versus secondary/tertiary care settings. We listed the 15 antibiotics that were ranked as the top-3 most frequently used antibiotics in at least 2 studies, grouped by Access and Watch. The ATC/DDD versions used in the studies were 2003 (Hadi 2008), 2011 (Dirga 2021, Pradipta 2015, Sholih 2019), 2013 (Kusuma 2016, Trisia 2020), 2017 (Herawati 2019, Narulita 2020), 2018 (Perdaka 2020, Rachmawati 2020), 2019 (Andriani 2020) or not reported (Kartika 2019, Pratama 2019, Mahmudah 2016, Massey 2021, Muliani 2021, Putri 2021, Susanto 2019, Wikantiananda 2019, Yulia 2017).

**Figure S1.** Forest plot of 20 reports on the defined daily dose (DDD) of total antibiotic use.

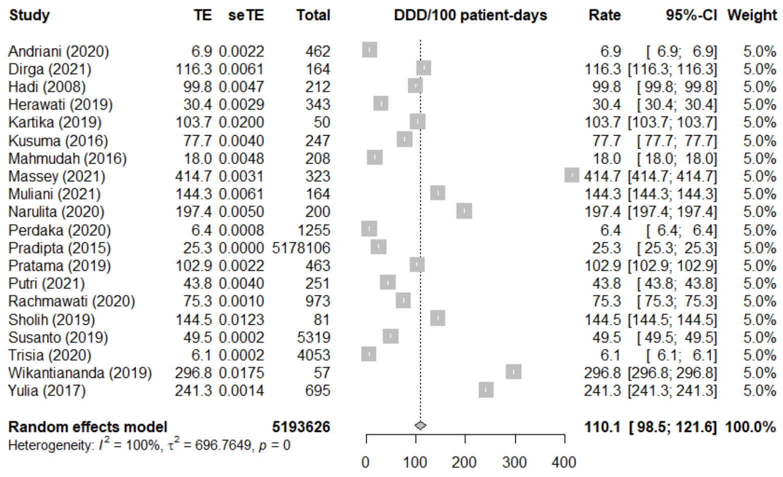

**Figure S2.** Results of leave-one-out sensitivity analysis for meta-analysis of the defined daily dose (DDD) of total antibiotic use.

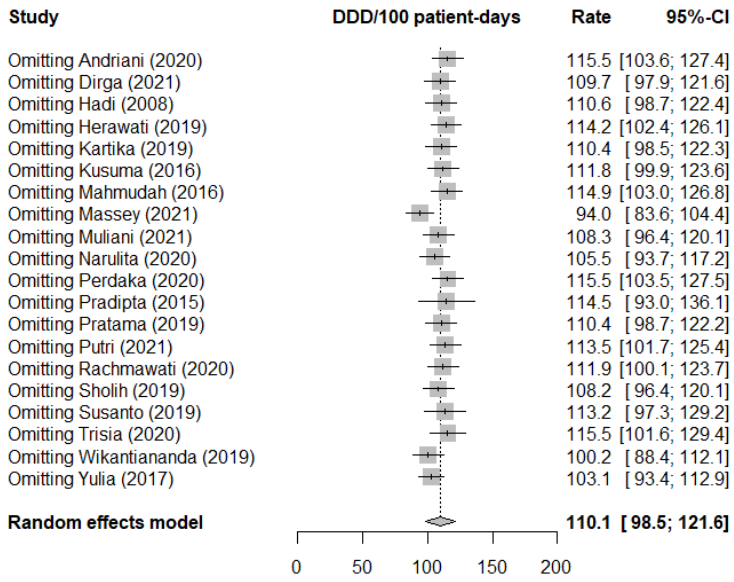

**Figure S3.** Funnel plot indicating evidence of publication bias (as shown by asymmetry) for meta-analysis of the defined daily dose (DDD) of total antibiotic use

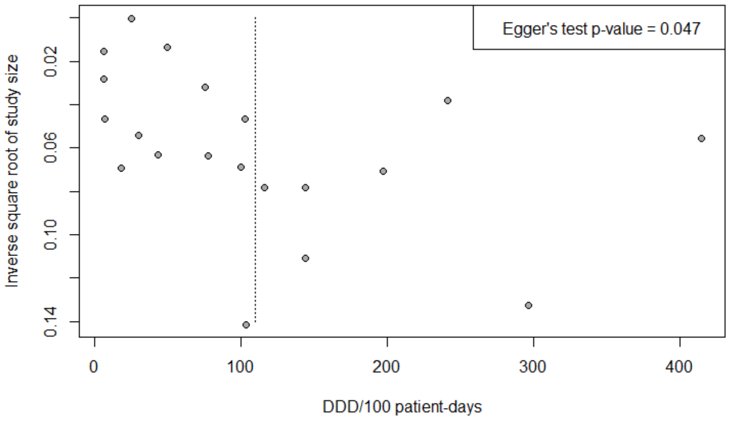

**Table S5**. Summary of studies on appropriateness of antibiotic prescribing (according to Gyssens method)

| First author | Year of study | Location | Healthcare level | Study population | No. of prescriptions | Classification | | | | | | | | | | | | |
|---|---|---|---|---|---|---|---|---|---|---|---|---|---|---|---|---|---|---|
| | | | | | | **iv** | **v** | **iv** | | | | **iii** | | **ii** | | | **i** | **Appropriate (0)** |
| | | | | | | | | **iva** | **ivb** | **ivc** | **ivd** | **iiia** | **iiib** | **iia** | **iib** | **iic** | | |
| Secondary/ tertiary healthcare | | | | | | | | | | | | | | | | | | |
| Aljufri[22] | 2017-2019 | Semarang | Tertiary | Pneumonia inpatients | 98 | 0.0 | 0.0 | 5.1 | 0.0 | - | 0.0 | 0.0 | 0.0 | 1.02 | 5.1 | 0.0 | 0.0 | 88.8 |
| Hanifah[24] | 2017 | Semarang | Secondary | Typhoid inpatients | 98 | 0.0 | 0.0 | 25.5 | 0.0 | 15.3 | 0.0 | 0.0 | 11.2 | 22.4 | 5.1 | 0.0 | 0.0 | 20.4 |
| Hardiana[25] | 2019 | Jakarta | Tertiary | Pneumonia inpatients | 88 | 0.0 | 0.0 | 7.9 | 0.0 | 2.3 | 0.0 | 5.7 | 10.2 | 0.0 | 0.0 | 0.0 | 0.0 | 73.9 |
| Ibrahim[26] | 2019 | Surabaya | Tertiary | Inpatients with infection | 84 | 36.9 | 11.9 | 6.0 | 0.0 | 0.0 | 0.0 | 0.0 | 0.0 | 0.0 | 1.2 | 0.0 | 0.0 | 44.1 |
| Inez[27] | 2018 | Tanjungpur a | Secondary | Paediatric inpatients | 63 | 4.8 | 82.5 | 11.1 | 0.0 | 0.0 | 0.0 | 0.0 | 1.6 | 0.0 | 0.0 | 0.0 | 0.0 | 0.0 |
| Kartika[5] | 2018 | Semarang | Secondary | Internal medicine inpatients (pre-intervention) | 50 | 0.0 | 46 | 12 | 2.0 | 0.0 | 2.0 | 0.0 | 2.0 | 6.0 | 0.0 | 2.0 | 0.0 | 28.0 |
| Maakh[28] | 2018 | Atambua | Secondary | Ob/gyn inpatients | 100 | 9.0 | 7.0 | 0.0 | 0.0 | 0.0 | 0.0 | 0.0 | 0.0 | 0.0 | 0.0 | 0.0 | 0.0 | 84.0 |
| Magdalena[29] | 2017 | Pekanbaru | Secondary | ICU inpatients | 307 | 0.0 | 2.6 | 6.5 | 0.0 | 0.0 | 4.9 | 10.1 | 1.6 | 3.9 | 2.6 | 0.0 | 0.0 | 67.8 |
| | | | | Medical ward inpatients | 570 | 0.0 | 5.3 | 9.1 | 0.0 | 0.0 | 5.4 | 4.2 | 1.8 | 3.7 | 1.8 | 1.4 | 0.0 | 67.4 |
| Muthoharoh[30] | 2017 | Pekalongan | Secondary | Surgical inpatients | 100 | 0.0 | 13.0 | 73.0 | 0.0 | 1.0 | 0.0 | 3.0 | 0.0 | 0.0 | 0.0 | 0.0 | 0.0 | 10.0 |
| Purwaningsih[31] | 2014-2015 | Semarang | Secondary | Paediatric inpatients | 385 | - | 8.6 | 22.1 | 0.0 | 20.0 | 1.6 | 0.0 | 0.0 | 44.4 | 37.7 | 0.0 | 0.0 | 23.9 |
| Rosdiana[32] | 2016 | Pekanbaru | Secondary | Inpatients (pre-intervention) | 92 | 0.0 | 27.2 | 35.9 | 0.0 | 2.2 | 0.0 | 0.0 | 0.0 | 1.1 | 0.0 | 0.0 | 0.0 | 33.7 |
| Setiawan[33] | 2016 | Surabaya | Tertiary | ICU inpatients | 110 | 31.8 | 7.3 | - | - | - | - | - | - | - | - | - | - | 52.7 |
| Sumiwi[34] | 2013 | Bandung | Secondary | Digestive surgery inpatients | 344 | - | 35.2 | 57.8 | 0.0 | 0.0 | 0.0 | 2.0 | 0.30 | 1.5 | 0.3 | 0.0 | 0.0 | 2.9 |

| First author | Year of study | Location | Healthcare level | Study population | No. of prescriptions | Classification | | | | | | | | | | | | |
|---|---|---|---|---|---|---|---|---|---|---|---|---|---|---|---|---|---|---|
| | | | | | | iv | v | iv | | | | iii | | ii | | | i | Appropriate (0) |
| | | | | | | | | iva | ivb | ivc | ivd | iiia | iiib | iia | iib | iic | | |
| Sutrisno[35] | 2011-2012 | Yogyakarta | Tertiary | Pneumonia inpatients | 57 | 0.0 | 2.0 | 5.9 | 0.0 | 1.0 | 0.0 | 7.9 | 6.9 | 2.0 | 6.9 | 17.8 | - | 49.5 |
| Waridiarto[36] | 2015 | Semarang | Tertiary | Orthopaedic inpatients | 150 | 1.3 | 42 | 0.0 | 0.0 | 0.0 | 8.0 | 3.3 | 0.0 | 0.0 | 0.0 | 0.0 | - | 45.3 |
| Yoanitha[37] | 2016 | Bandung | Tertiary | Ob/gyn inpatients | 236 | 0.0 | 40.3 | - | - | - | - | 0.0 | 0.0 | 0.0 | 0.0 | 0.0 | 0.0 | 22.0 |
| Primary healthcare | | | | | | | | | | | | | | | | | | |
| Adani[21] | 2014 | Semarang | Primary | Paediatric inpatients | 173 | 0.0 | 62.4 | 1.7 | 0.0 | 0.0 | 0.0 | 0.0 | 0.0 | 0.0 | 0.0 | 5.2 | - | 30.6 |
| Anggraini[23] | 2014 | Pontianak | Primary | Typhoid inpatients | 62 | 0.0 | 4.8 | 3.2 | 0.0 | 0.0 | 0.0 | 0.0 | 16.1 | 0.0 | 0.0 | 0.0 | 6.5 | 69.4 |

Abbreviations: -, not available; ICU, intensive care unit; PICU, paediatric intensive care unit; Ob/Gyn, Obstetrics and gynaecology.

The table summarizes 18 studies on the appropriateness of antibiotic prescriptions based on the Gyssens method. Gyssens evaluates the quality of each antibiotic prescription by sequentially categorizing 6 indicators from vi to i**:** vi=insufficient data; v=antibiotic is not indicated; iv=alternative antibiotic is available that is more effective (iva), less toxic (ivb), less costly (ivc); has narrower spectrum (ivd); iii=inappropriate duration: too long (iiia) or too short (iiib); ii=incorrect: dose (iia), interval (iib), route (iic); i=incorrect timing.

**Table S6.** Summary of studies on appropriateness of antibiotic prescribing (according to reference guidelines)

| First author | Year of study | Location | Health care level | Study population | No. of prescriptions | Appropriate (%) | | | | | | | | Reference guidelines |
|---|---|---|---|---|---|---|---|---|---|---|---|---|---|---|
| | | | | | | **No contra-indication/ allergy label** | **Indication** | **Drug choice** | **Dose** | **Dosing frequency** | **Duration** | **Administration route** | **Overall rational use** | |
| Secondary/tertiary health care | | | | | | | | | | | | | | |
| Anggraini[39] | 2020 | Jakarta | Secondary | Pre-surgical inpatients | 837 | - | - | 0.5 | 0.5 | - | - | 0.5 | 0.0 | IN |
| Bakhtiar[40] | 2018 | Sorong | Secondary | Post-surgical inpatients | 83 | 94 | 100 | 100 | 100 | - | 92.8 | 100 | 79.5[a] | IN |
| Bestari[42] | 2016 | Central Java | Tertiary | Paediatric pneumonia inpatients | 90 | 100 | 100 | 72.2 | 9.2 | - | - | - | 8.9 | NH |
| Dania[43] | 2014 | Yogyakarta | Secondary | Ob/gyn inpatients (therapy) | 59 | - | - | - | - | - | 100 | - | - | IHB |
| Dewi[44] | 2017 | Jakarta | Tertiary | Adult ICU inpatients with sepsis or septic shock | 60 | - | - | 66.7 | 48.3 | - | - | - | - | I |
| Dirga[2] | 2017 | Lampung | Secondary | Internal medicine inpatients | 168 | 100 | 98.8 | 89.3 | 52.9 | - | - | - | - | INBH |
| Elvina[47] | 2016 | Jakarta | Secondary | Adult inpatients with pneumonia | 96 | - | - | 86.5 | 91.7 | - | 74 | - | - | INB |
| Fithria[49] | 2013 | Semarang | Secondary | Paediatric inpatients with acute diarrhoea | 54 | - | - | 0 | - | - | - | - | - | IB |
| Grassella[50] | 2018 | Pontianak | Secondary | Paediatric ARI outpatients | 340 | 99.7 | 91.7 | 72.9 | 72.6 | 96.5 | 56.8 | 100 | - | NBW |
| Harartasyahrani[51] | 2020 | Prabumulih | Secondary | Pre-surgical inpatients | 119 | - | - | 92.4 | 86.4 | 7.3 | 40 | 100 | 6.7 | IN |
| Herlina[53] | 2017 | Mataram | Secondary | UTI inpatients | 105 | - | - | 100 | 100 | 99.1 | 88.6 | - | - | I |
| Islam[54] | 2014 | Jakarta | Tertiary | Pneumonia inpatients | 139 | - | - | 5.7 | 88.5 | - | 60.4 | - | - | INBW |
| Kaparang[56] | 2013 | Manado | Tertiary | Paediatric pneumonia inpatients | 112 | 100 | 100 | 100 | 91.1 | - | 88.4 | - | - | NHB |
| Kurniawati[57] | 2019 | Yogyakarta | Secondary | UTI inpatients | 61 | - | - | 100 | 88.5 | 98.4 | 88.5 | 100 | - | IB |
| Limato[58] | 2019 | Jakarta | Secondary / Tertiary | Hospital-wide inpatients | 915 | - | - | 52.2 | - | - | - | - | - | HN |
| Nawakasari[59] | 2017 | Klaten | Tertiary | UTI inpatients | 76 | 100 | 100 | 96.1 | 27.6 | - | - | - | - | INBH |
| Octavia[61] | 2018 | Lamongan | Secondary | Ob/gyn inpatients (prophylaxis) | 54 | - | - | 98.1 | 100 | 100 | - | 100 | - | I |
| Ofisya[62] | 2019 | Pontianak | Secondary | Pneumonia inpatients | 86 | - | 100 | 93 | 68.6 | - | 62.8 | - | - | IN |
| Oktaviani[63] | 2014-2015 | Riau | Secondary | Ob/gyn inpatients (prophylaxis) | 140 | - | - | 34.3 | 65.7 | 100 | 72.9 | 100 | - | INB |
| Rusdiana[65] | 2014 | Tangerang | Secondary | Ob/gyn inpatients (prophylaxis) | 256 | - | - | 0.0 | 7.8 | 0 | - | 100 | - | B |
| Zazuli[67] | 2009 | Bandung | Secondary | Pre- and post-surgical inpatients | 1290 | - | 99.6 | - | 97.2 | - | - | - | - | IB |
| Primary health care | | | | | | | | | | | | | | |
| Andrajati[38] | 2012 | Depok | Primary | Outpatients with various indications | 392 | - | - | 87.2 | - | 98.2 | 59.4 | - | 43.9 | N |
| Benua[41] | 2018 | Poso | Primary | ARI outpatients | 126 | - | 100 | - | 80.9 | 80.9 | 100 | 100 | - | N |

| First author | Year of study | Location | Health care level | Study population | No. of prescriptions | Appropriate (%) | | | | | | | | Reference guidelines |
|---|---|---|---|---|---|---|---|---|---|---|---|---|---|---|
| | | | | | | **No contra-indication/ allergy label** | **Indication** | **Drug choice** | **Dose** | **Dosing frequency** | **Duration** | **Administration route** | **Overall rational use** | |
| Dewi[45] | 2018 | Jambi | Primary | Paediatric ARI outpatients | 70 | 98.5 | 100 | 54.2 | - | 48.5 | 1.4 | - | - | N |
| Dewi[46] | 2019 | Jambi | Primary | Toddler ARI outpatients | 51 | 100 | 100 | - | 100 | - | 0 | - | - | N |
| Fakhrunnisa[48] | 2018 | Tegal | Primary | ARI outpatients | 632 | - | - | 78.0 | 90.5 | 98.2 | 33.7 | - | 1.7 | N |
| Hasyul[52] | 2017 | Garut | Primary | Typhoid outpatients | 705 | - | 96.9 | 58.3 | 63.3 | - | 49.8 | - | - | N |
| Jamiati[55] | 2017 | Aceh | Primary | Outpatients | 86 | - | 54.6 | - | 60.5 | 83.7 | 26.7 | - | - | N |
| Nyoman[60] | 2015 | Bandung | Primary | Paediatric ARI outpatients | 425 | - | 100 | 96.5 | 42.2 | - | 52.8 | - | - | I |
| Ovikariani[64] | 2019 | Semarang | Primary | ARI outpatients | 79 | 100 | 23.0 | 23.0 | 82.3 | - | - | - | - | N |
| Sugiarti[66] | 2014 | Malang | Primary | Paediatric ARI outpatients | 120 | - | 24.2 | 100 | 8.9 | - | - | - | - | N |

Abbreviations: ARI, acute respiratory infection; -, not available; UTI, urinary tract infection; ICU, intensive care unit; Ob/Gyn, obstetrics and gynaecology; I, international; N, national; H, hospital; B, book; W, website

The table summarizes 31 studies on appropriateness of antibiotic prescribing, based on antibiotic prescribing reference guidelines (as provided in the article), divided by primary and secondary/tertiary care settings. Appropriateness was defined by the following indicators: indication, drug choice, dose, frequency/interval, duration, and route of administration. An antibiotic prescription was classified as rational use if all indicators were scored as compliant with the reference guideline by the authors.

**Figure S4.** Forest plot showing results of meta-analysis on the overall appropriateness of antibiotic prescribing according to Gyssens method (18 reports)

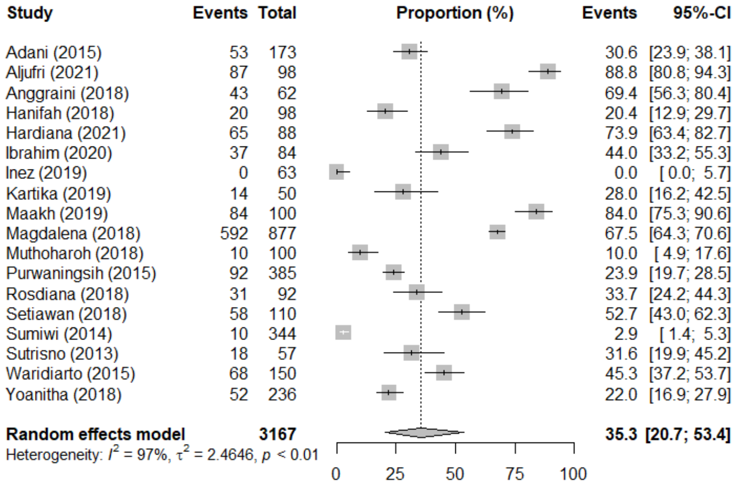

**Figure S5.** Results of leave-one-out sensitivity analysis for meta-analysis on the overall appropriateness of antibiotic prescribing according to Gyssens method

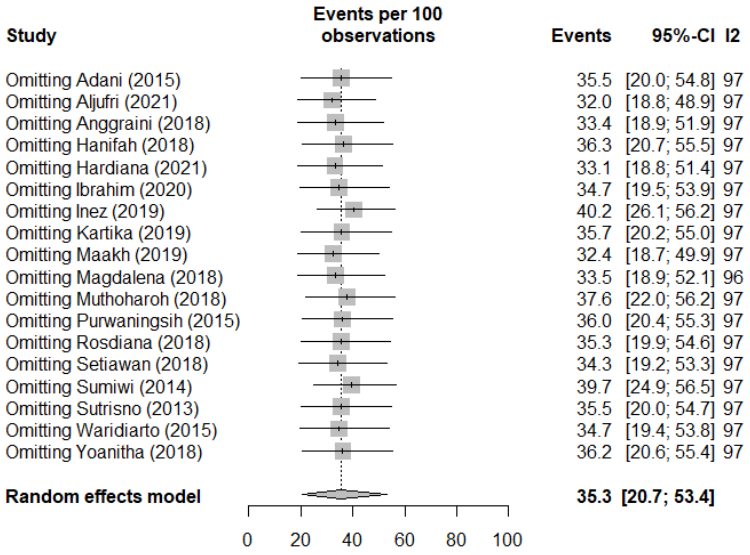

**Figure S6.** Results of sensitivity analysis for meta-analysis on the overall appropriateness of antibiotic prescribing according to Gyssens method by using alternative methods other than generalized linear mixed model (GLMM) method.

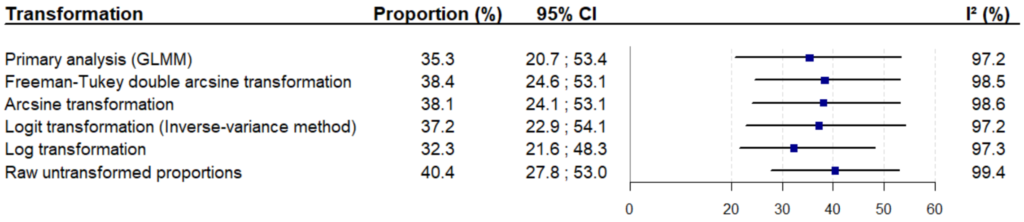

**Figure S7.** Funnel plot for the meta-analysis on the overall appropriateness of antibiotic prescribing according to Gyssens method. The relatively symmetrical funnel plot and the non-significant Egger's test indicate that no publication bias is detected.

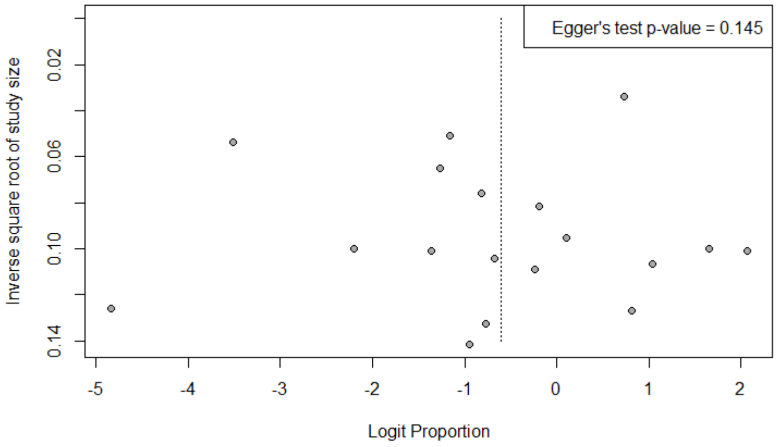

**Figure S8.** Summary forest plot of 19 reports on the appropriateness of antibiotic prescribing according to the "duration"  indicator in the reference guidelines

| Variable | N | Events | Total | Proportion (%) | 95% CI | I² (%) | Egger's test p-value |
|---|---|---|---|---|---|---|---|
| **Health care setting** | | | | | | | |
| Primary care | 8 | 1016 | 2180 | 34.5 | 4.3 ; 86.1 | 93.5 | NA |
| Hospital | 11 | 930 | 1331 | 80.0 | 66.0 ; 89.2 | 91.9 | NA |
| **Study location** | | | | | | | |
| Java | 8 | 1134 | 2202 | 70.9 | 47.5 ; 86.8 | 94.8 | NA |
| Non-Java | 11 | 812 | 1309 | 58.5 | 18.1 ; 90.0 | 94.0 | NA |
| **Year of publication** | | | | | | | |
| <2016 | 6 | 802 | 1267 | 79.1 | 53.8 ; 92.5 | 89.7 | NA |
| ≥2016 | 13 | 1144 | 2244 | 56.1 | 22.1 ; 85.2 | 95.1 | NA |
| **Age groups** | | | | | | | |
| Adults | 8 | 584 | 793 | 83.2 | 65.3 ; 92.9 | 92.1 | NA |
| Children | 5 | 518 | 998 | 17.9 | 1.2 ; 79.4 | 93.1 | NA |
| Mixed | 6 | 844 | 1720 | 67.8 | 26.4 ; 92.5 | 94.6 | NA |
| **Total** | **19** | **1946** | **3511** | **65.8** | **38.6 ; 85.5** | **94.6** | **0.074** |

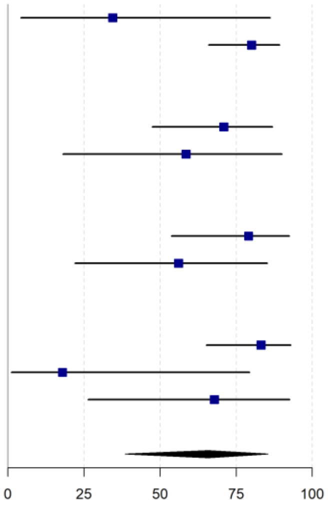

**Figure S9.** Summary forest plot of 27 reports on the appropriateness of antibiotic prescribing according to the “drug choice” indicator in the reference guidelines

| Variable | N | Events | Total | Proportion (%) | 95% CI | I² (%) | Egger's test p-value |
|---|---|---|---|---|---|---|---|
| **Health care setting** | | | | | | | |
| Primary care | 7 | 1782 | 2372 | 82.6 | 50.5 ; 95.7 | 97.7 | NA |
| Hospital | 20 | 1865 | 3850 | 84.9 | 38.5 ; 98.0 | 96.0 | NA |
| **Study location** | | | | | | | |
| Java | 18 | 2668 | 4999 | 70.0 | 23.2 ; 94.7 | 97.2 | NA |
| Non-Java | 9 | 979 | 1223 | 95.2 | 73.9 ; 99.3 | 94.7 | NA |
| **Year of publication** | | | | | | | |
| <2016 | 9 | 1071 | 1668 | 80.7 | 1.5 ; 99.9 | 97.4 | NA |
| ≥2016 | 18 | 2576 | 4554 | 85.7 | 61.7 ; 95.7 | 96.2 | NA |
| **Age groups** | | | | | | | |
| Adults | 14 | 881 | 2253 | 89.3 | 33.1 ; 99.3 | 96.7 | NA |
| Children | 7 | 966 | 1182 | 89.2 | 16.5 ; 99.7 | 93.0 | NA |
| Mixed | 6 | 1800 | 2787 | 69.2 | 45.5 ; 85.8 | 98.0 | NA |
| **Total** | **27** | **3647** | **6222** | **84.3** | **52.3 ; 96.3** | **96.6** | **0.305** |

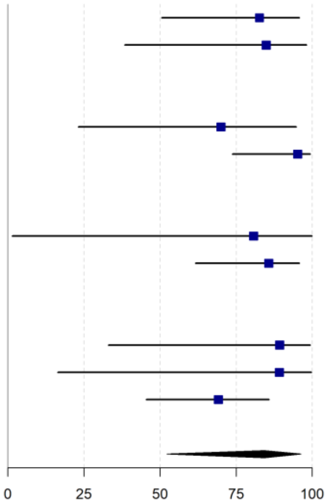

**Figure S10.** Summary forest plot of 27 reports on the appropriateness of antibiotic prescribing according to the “dose” indicator in the reference guidelines

| Variable | N | Events | Total | Proportion (%) | 95% CI | I² (%) | Egger's test p-value |
|---|---|---|---|---|---|---|---|
| **Health care setting** | | | | | | | |
| Primary care | 8 | 1274 | 1888 | 72.9 | 42.8 ; 90.6 | 97.8 | NA |
| Hospital | 19 | 2493 | 4044 | 82.9 | 82.5 ; 83.2 | 97.8 | NA |
| **Study location** | | | | | | | |
| Java | 16 | 2690 | 4525 | 67.2 | 32.3 ; 89.8 | 98.6 | NA |
| Non-Java | 11 | 1077 | 1407 | 90.0 | 71.1 ; 97.1 | 85.3 | NA |
| **Year of publication** | | | | | | | |
| <2016 | 8 | 1748 | 2418 | 75.2 | 32.6 ; 95.0 | 98.9 | NA |
| ≥2016 | 19 | 2019 | 3514 | 82.0 | 54.0 ; 94.6 | 96.0 | NA |
| **Age groups** | | | | | | | |
| Adults | 15 | 2079 | 3440 | 88.1 | 52.0 ; 98.1 | 98.1 | NA |
| Children | 6 | 595 | 1085 | 64.3 | 16.7 ; 94.2 | 97.4 | NA |
| Mixed | 6 | 1093 | 1407 | 75.9 | 65.7 ; 83.8 | 95.3 | NA |
| **Total** | **27** | **3767** | **5932** | **80.0** | **57.3 ; 92.3** | **97.7** | **0.732** |

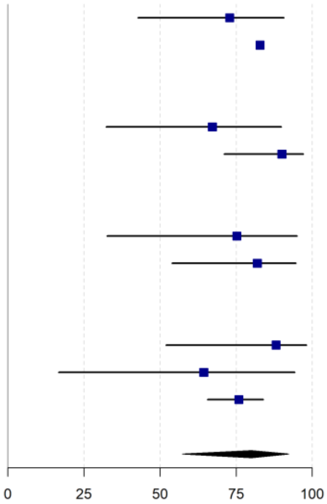

**Figure S11.** Summary forest plot of 6 reports on the appropriateness of antibiotic prescribing according to the “overall rational use” indicator in the reference guidelines

| Variable | N | Events | Total | Proportion (%) | 95% CI | I² (%) | Egger's test p-value |
|---|---|---|---|---|---|---|---|
| **Healthcare setting** | | | | | | | |
| Primary care | 2 | 183 | 1024 | 10.5 | 0.8 ; 62.6 | 99.3 | NA |
| Hospital | 4 | 82 | 524 | 6.1 | 0.2 ; 63.2 | 97.1 | NA |
| **Study location** | | | | | | | |
| Java | 4 | 191 | 1346 | 3.3 | 0.3 ; 32.0 | 98.1 | NA |
| Non-Java | 2 | 74 | 202 | 34.5 | 3.1 ; 89.8 | 98.7 | NA |
| **Year of publication** | | | | | | | |
| <2016 | 1 | 172 | 392 | 43.9 | 39.0 ; 48.8 | NA | NA |
| ≥2016 | 5 | 93 | 1156 | 5.0 | 4.7 ; 5.3 | 97.9 | NA |
| **Age groups** | | | | | | | |
| Adults | 3 | 74 | 434 | 4.2 | 0.0 ; 86.6 | 97.4 | NA |
| Children | 1 | 8 | 90 | 8.9 | 4.5 ; 16.8 | NA | NA |
| Mixed | 2 | 183 | 1024 | 10.5 | 0.8 ; 62.6 | 99.3 | NA |
| **Total** | **6** | **265** | **1548** | **8.0** | **1.0 ; 43.6** | **97.9** | **NA** |

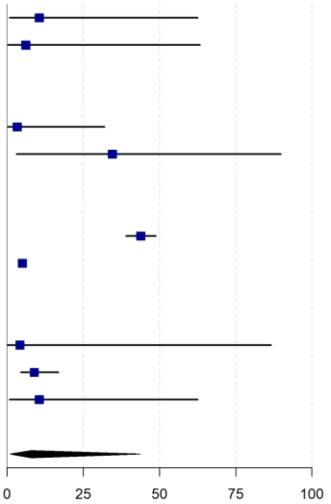

**Figure S12.** Summary forest plot of 9 reports on the appropriateness of antibiotic prescribing according to the “no contraindication/allergy label” indicator in the reference guidelines

| Variable | N | Events | Total | Proportion (%) | 95% CI | I² (%) | Egger's test p-value |
|---|---|---|---|---|---|---|---|
| **Health care setting** | | | | | | | |
| Primary care | 3 | 199 | 200 | 99.5 | 96.5 ; 99.9 | 35.9 | NA |
| Hospital | 6 | 863 | 869 | 99.9 | 96.4 ; 100.0 | 0.0 | NA |
| **Study location** | | | | | | | |
| Java | 3 | 245 | 245 | 100.0 | 0.0 ; 100.0 | 0.0 | NA |
| Non-Java | 6 | 817 | 824 | 99.6 | 97.0 ; 99.9 | 41.5 | NA |
| **Year of publication** | | | | | | | |
| <2016 | 1 | 112 | 112 | 100.0 | 96.8 ; 100.0 | NA | NA |
| ≥2016 | 8 | 950 | 957 | 99.7 | 97.7 ; 100.0 | 18.1 | NA |
| **Age groups** | | | | | | | |
| Adults | 3 | 322 | 327 | 99.8 | 70.0 ; 100.0 | 0.0 | NA |
| Children | 5 | 661 | 663 | 99.7 | 98.8 ; 99.9 | 0.0 | NA |
| Mixed | 1 | 79 | 79 | 100.0 | 95.4 ; 100.0 | NA | NA |
| **Total** | **9** | **1062** | **1069** | **99.8** | **98.0 ; 100.0** | **6.4** | **NA** |

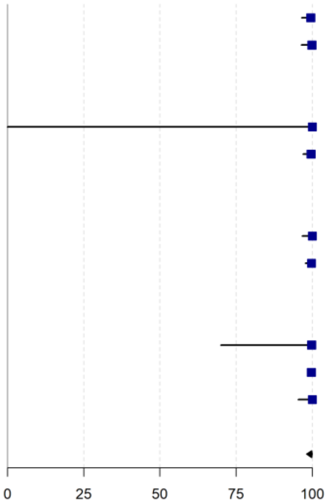

**Figure S13.** Summary forest plot of 16 reports on the appropriateness of antibiotic prescribing according to the “indications” indicator in the reference guidelines

| Variable | N | Events | Total | Proportion (%) | 95% CI | $I^2$ (%) | Egger's test p-value |
|---|---|---|---|---|---|---|---|
| **Health care setting** | | | | | | | |
| Primary care | 8 | 1449 | 1662 | 98.9 | 60.9 ; 100.0 | 97.6 | NA |
| Hospital | 8 | 2185 | 2220 | 99.7 | 97.4 ; 100.0 | 84.5 | NA |
| **Study location** | | | | | | | |
| Java | 7 | 2606 | 2785 | 99.0 | 70.1 ; 100.0 | 98.5 | NA |
| Non-Java | 9 | 1028 | 1097 | 99.8 | 99.8 ; 99.8 | 89.3 | NA |
| **Year of publication** | | | | | | | |
| <2016 | 4 | 1851 | 1947 | 99.7 | 53.2 ; 100.0 | 98.4 | NA |
| ≥2016 | 12 | 1783 | 1935 | 99.5 | 92.8 ; 100.0 | 95.7 | NA |
| **Age groups** | | | | | | | |
| Adults | 4 | 1610 | 1617 | 99.6 | 99.1 ; 99.8 | 0.0 | NA |
| Children | 7 | 1089 | 1208 | 100.0 | 76.2 ; 100.0 | 96.0 | NA |
| Mixed | 5 | 935 | 1057 | 96.6 | 41.2 ; 99.9 | 98.1 | NA |
| **Total** | **16** | **3634** | **3882** | **99.6** | **95.2 ; 100.0** | **96.9** | **0.092** |

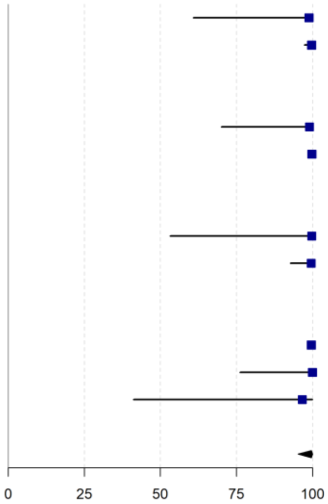

**Figure S14.** Summary forest plot of 13 reports on the appropriateness of antibiotic prescribing according to the “dosing frequency” indicator in the reference guidelines

| Variable | N | Events | Total | Proportion (%) | 95% CI | $I^2$ (%) | Egger's test p-value |
|---|---|---|---|---|---|---|---|
| **Health care setting** | | | | | | | |
| Primary care | 5 | 1214 | 1306 | 90.3 | 69.4 ; 97.4 | 97.3 | NA |
| Hospital | 8 | 753 | 1125 | 97.9 | 18.2 ; 100.0 | 96.0 | NA |
| **Study location** | | | | | | | |
| Java | 6 | 1179 | 1454 | 97.7 | 12.9 ; 100.0 | 0.0 | NA |
| Non-Java | 7 | 788 | 977 | 89.5 | 48.7 ; 98.7 | 96.9 | NA |
| **Year of publication** | | | | | | | |
| <2016 | 4 | 584 | 847 | 100.0 | 0.0 ; 100.0 | 0.0 | NA |
| ≥2016 | 9 | 1383 | 1584 | 92.4 | 68.1 ; 98.6 | 97.2 | NA |
| **Age groups** | | | | | | | |
| Adults | 7 | 425 | 785 | 99.3 | 2.1 ; 100.0 | 92.0 | NA |
| Children | 2 | 362 | 410 | 83.7 | 32.7 ; 98.2 | 98.7 | NA |
| Mixed | 4 | 1180 | 1236 | 94.3 | 82.7 ; 98.3 | 95.8 | NA |
| **Total** | **13** | **1967** | **2431** | **94.2** | **59.3 ; 99.5** | **96.3** | **0.580** |

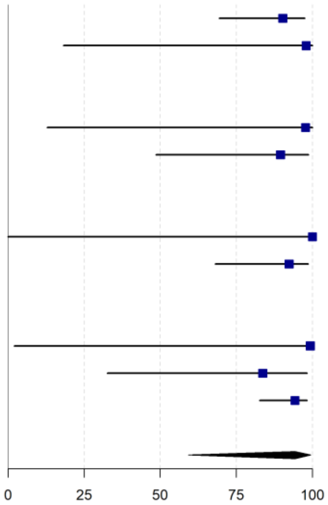

**Figure S15.** Summary forest plot of 9 reports on the appropriateness of antibiotic prescribing according to the “administration route” indicator in the reference guidelines

| Variable | N | Events | Total | Proportion (%) | 95% CI | I² (%) | Egger's test p-value |
|---|---|---|---|---|---|---|---|
| **Health care setting** | | | | | | | |
| Primary care | 1 | 126 | 126 | 100.0 | 97.1 ; 100.0 | NA | NA |
| Hospital | 8 | 1048 | 1881 | 100.0 | 95.5 ; 100.0 | 0.0 | NA |
| **Study location** | | | | | | | |
| Java | 4 | 375 | 1208 | 100.0 | 42.6 ; 100.0 | 0.0 | NA |
| Non-Java | 5 | 799 | 799 | 100.0 | 0.0 ; 100.0 | 0.0 | NA |
| **Year of publication** | | | | | | | |
| <2016 | 2 | 396 | 396 | 100.0 | 0.0 ; 100.0 | 0.0 | NA |
| ≥2016 | 7 | 778 | 1611 | 100.0 | 92.8 ; 100.0 | 0.0 | NA |
| **Age groups** | | | | | | | |
| Adults | 7 | 708 | 1541 | 100.0 | 93.1 ; 100.0 | 0.0 | NA |
| Children | 1 | 340 | 340 | 100.0 | 98.9 ; 100.0 | NA | NA |
| Mixed | 1 | 126 | 126 | 100.0 | 97.1 ; 100.0 | NA | NA |
| **Total** | **9** | **1174** | **2007** | **100.0** | **97.2 ; 100.0** | **0.0** | **NA** |

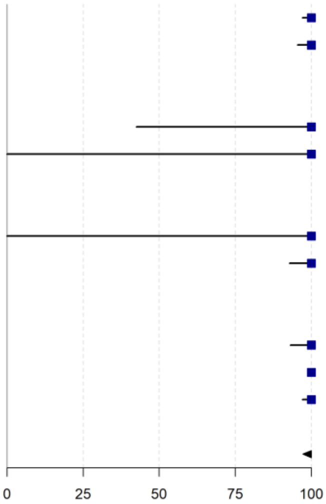

**Table S7.** Summary of studies on knowledge, attitudes and perceptions regarding antibiotic use

| First Author | Year of study | Location | Study population | No respondents | Questionnaire | AMR awareness | Antibiotic use knowledge | Self-medication | Where to buy antibiotic | Advice to take antibiotic/ to practice self-medication | Reasons for self-medication | Adherence to treatment | Antibiotic stewardship |
|---|---|---|---|---|---|---|---|---|---|---|---|---|---|
| Community | | | | | | 8 | 16 | 11 | 9 | 10 | 6 | 2 | 1 |
| Artini[76] | 2014-2015 | Denpasar, Bali | Community respondents: medical and non-medical university students | 240 | Developed by the authors | | • | • | • | • | • | | |
| Djawaria[78] | 2014-2015 | Surabaya, Easy Java | Community respondents at pharmacy | 267 | Existing questionnaire (Djawaria, 2018) | | | • | • | • | | | |
| Fatmah[79] | 2018 | Mataram, West Nusa Tenggara | Community: medical and non-medical university students | 400 | Based on previous studies | | • | • | • | | | | |
| Fernandez[80] | 2012 | Manggarai Barat, East Nusa Tenggara | Community respondents at pharmacies | 108 | Developed by the authors | | • | • | • | • | • | | |
| Fimanggara[81] | 2013 | Jatinangor, West Java | Community respondents: non-medical college students | 250 | Modified from previous study | • | • | | | | | | |
| Fitriah[82] | 2019 | Banjarbaru, South Kalimantan | Community respondents | 380 | Developed by the authors | | • | | | | | | |
| Hamid[83] | 2019 | Pangkajene and Kepulauan, South Sulawesi | Community respondents: teachers in public schools | 236 | Developed by the authors | | | • | • | • | • | | |
| Insany[84] | 2014 | Bandung, West Java | Outpatient respondents from primary health centres and pharmacies | 508 | Based on previous studies | | | • | | | | | |
| Kondoj[85] | 2019-2020 | Manado, North Sulawesi | Community respondents | 290 | Developed by the authors | • | • | • | | • | | | |
| Kristina[86] | 2018 | Yogyakarta, DIY | Patients at outpatient clinics and pharmacies | 268 | Modified from previous study | • | • | | | • | | | |
| Kurniawan[87] | 2015 | Manado, North Sulawesi | Community respondents at primary health centre | 400 | Modified from previous studies | | • | • | • | • | • | | |
| Novelni[88] | 2019 | Padang, West Sumatra | Community respondents | 100 | Developed by the authors | • | • | | | | | | |
| Nuraini[89] | 2017-2018 | Bangkalan, East Java | Outpatients at secondary hospital | 103 | Developed by the authors | | • | | | | | • | |

| First Author | Year of study | Location | Study population | No respondents | Questionnaire | AMR awareness | Antibiotic use knowledge | Self-medication | Where to buy antibiotic | Advice to take antibiotic/ to practice self-medication | Reasons for self-medication | Adherence to treatment | Antibiotic stewardship |
|---|---|---|---|---|---|---|---|---|---|---|---|---|---|
| Pratama[90] | 2016 | Jember, East Java | Community respondents: non-medical university students | 324 | Developed by the authors | | | • | • | | | | |
| Salsabila[91] | 2019 | Yogyakarta, DIY | Community respondents in rural and urban areas | 125 | Based on previous studies | • | • | | | • | | | |
| Siahaan[92] | 2015 | West Java, DKI Jakarta, Southeast Sulawesi | Community respondents: household members | 1271 | Developed by the authors | | | | | | | • | |
| Tandjung[94] | 2020-2021 | Manado, North Sulawesi | Community respondents | 323 | Developed by the authors | | • | • | | | • | | |
| WHO[95] | 2015 | National | Community respondents: household members | 1027 | Developed by specialized research agency, in collaboration with WHO | • | • | | • | • | | | • |
| Widayati[96] | 2010 | Yogyakarta, DIY | Community respondents: household members | 559 | Based on previous studies in Asia | | | • | • | • | • | | |
| Widayati[97] | 2010 | Yogyakarta, DIY | Community respondents: household members | 559 | Based on previous studies in Asia | • | • | | | | | | |
| Yulia[98] | 2019 | Bukittinggi, West Sumatra | Community respondents | 100 | Developed by the authors | • | • | | | | | | |
| Yuliani[99] | 2014 | Kupang, East Nusa Tenggara | Community respondents | 100 | Developed by the authors | | • | | | | | | |
| Healthcare provider | | | | | | Antibiotic dispensing (3) | | | Antibiotic use knowledge (1) | | The use of antibiotic guidelines (1) | | |
| Asvinigita[77] | 2019 | Yogyakarta, DIY | Pharmacists | 250 | Developed by the authors | • | | | | | | | |
| Siswati[93] | 2000 | Padang, West Sumatra | Health care workers | 110 | Developed by the authors | • | | | • | | • | | |
| Zhang[100] | 2018 | National | General practitioners and paediatricians | 100 | Modified from existing questionnaire developed by World Gastroenterology Organization | • | | | | | | | |

Abbreviations: AMR, antimicrobial resistance; DIY, Daerah Istimewa Yogyakarta; DKI, Daerah Khusus Ibukota; WHO, World Health Organization

The table summarizes 25 surveys in the domain attitudes and perceptions on antibiotic use, conducted among health care providers and communities, here organised by key emerging themes. Further details are provided in Table S5.

**Table S8**. Summary of findings of the knowledge, attitudes and practice surveys

| First author | Year of publication | Year of study | Location | Study population | No. of population | Themes | Summary of findings |
|---|---|---|---|---|---|---|---|
| **Community** | | | | | | | |
| **Artini**[76] | 2016 | 2014-2015 | Denpasar, Bali | Community respondents: medical and non-medical university students | 240 | Antibiotic use knowledge | Both medical and non-medical students were aware that antibiotic use can drive antibiotic resistance (91.7% and 84.2%) and antibiotic cannot treat all disease (90% and 92.5%). Medical students had better knowledge than non-medical student that antibiotics aim to treat bacterial infection (90.8% versus 76.7%) and not to treat viral infection (64.2% versus 49.2%). |
| | | | | | | Self-medication | Medical students were more likely to self-medicate (67.5%) than non-medical students (19.2%). |
| | | | | | | Where to buy antibiotic | >90% at the pharmacy, <10% at kiosks. |
| | | | | | | Advice to take antibiotic/ to practice self-medication | Past experience (36.5%), family (36.5%), and pharmacist (27%). |
| | | | | | | Reasons for self-medication | Self-medication was practical (77.9%) and cheaper (13.5%), doctor is far (8.6%). |
| **Djawaria**[78] | 2018 | 2014-2015 | Surabaya, East Java | Community respondents buying antibiotics without a prescription at pharmacy | 267 | Self-medication | Majority (76%) seldom self-medicate, and 12% purchased antibiotics using a previous doctor prescription. |
| | | | | | | Where to buy antibiotic | 51.3% at the pharmacy (51.3%), 9% from family and friends; others obtained antibiotics from kiosks, previous prescriptions, or unspecified. |
| | | | | | | Advice to take antibiotic/ to practice self-medication | Friends or family who work in the health sector (24%), family (21%), pharmacist (11%), self (6%), or friends who do not work in health sector (4%). |
| **Fatmah**[79] | 2 019 | 2018 | Mataram, West Nusa Tenggara | Community respondents: medical and non-medical university students | 400 | Antibiotic use knowledge | 67% of respondents used antibiotics for upper respiratory tract infection for 1-3 days as recommended by health care workers. |
| | | | | | | Self-medication | 47.5% self-medicated because it was easy to get, and many places sell antibiotics without prescriptions. Many were recommended to self-medicate by physicians or pharmacists. |
| | | | | | | Where to buy antibiotic | 88.1% respondents bought antibiotics from the pharmacy, 6.6% used the leftover antibiotics, 3.2% bought at kiosks, and 2.1% purchased at drugstores. |
| **Fernandez**[80] | 2013 | 2012 | Manggarai Barat, East Nusa Tenggara | Community respondents at pharmacies | 108 | Antibiotic use knowledge | 88% knew that antibiotics treat bacterial infection; 60-65% knew that the antibiotic course must be completed, cannot treat headache and can incur side-effects when used inappropriately. |
| | | | | | | Self-medication | All participants (100%) ever used antibiotics without a prescription. Most purchased the suggested full-course (84.2%) and kept antibiotic at home (65.7%). |

| First author | Year of publication | Year of study | Location | Study population | No. of population | Themes | Summary of findings |
|---|---|---|---|---|---|---|---|
| | | | | | | Where to buy antibiotic | At the pharmacy (87%), got it from family (12%) or friends (1%). |
| | | | | | | Advice to take antibiotic/ to practice self-medication | Doctor (40%), pharmacist (28%), self (18%), friends/family (8%), or brochure (6%). |
| | | | | | | Reasons for self-medication | 44.4% reported that a doctor's visit is too expensive, and 77% used the same antibiotic because it previously resulted in clinical improvement. |
| **Fimanggara**[81] | 2016 | 2013 | Jatinangor, West Java | Community respondents: non-medical students | 250 | AMR awareness and antibiotic use knowledge | Knowledge level was poor for 56.4%, moderate for 26.4% and good for 17.2%. Misconceptions were related to taking antibiotics for common cold, taking leftover antibiotics, and antibiotics are effective to treat the same symptoms as a previous infection and to treat inflammation. Good knowledge was shown on overuse antibiotic can cause AMR, antibiotics kill bacteria, and antibiotic should be used in according with instructions. |
| **Fitriah**[82] | 2021 | 2019 | Banjarbaru | Community respondents | 380 | Antibiotic use knowledge | 58.2% respondents had a good level of knowledge about antibiotic use. 98.4% had a positive attitude towards antibiotic use. Higher education ($p$=0.007) and higher income ($p$=0.038) was correlated with better antibiotic use knowledge. |
| **Hamid**[83] | 2020 | 2019 | Pangkajene and Kepulauan, South Sulawesi | Community respondents: teachers in public schools | 236 | Self-medication | 78.4% respondents had ever used antibiotics without prescription, and 53% kept antibiotics at home for future use. |
| | | | | | | Where to buy antibiotic | 78.9% at the pharmacy, and 21.1% at kiosks. |
| | | | | | | Advice to take antibiotic/ to practice self-medication | The highest source of information to self-medicate was relatives (38.4%), followed by pharmacists (30.3%), friends (12.4%), internet (12.4%), books (2.7%), kiosk staffs (1.6%), nurses (1.1%), midwives (0.5%), and television (0.5%). |
| | | | | | | Reasons for self-medication | 74.1% stated that previous good experience was the reason to self-medicate, and the rest mentioned no influence of previous experience. |
| **Insany**[84] | 2015 | 2014 | Bandung, West Java | Outpatients from 43 primary health centres and 8 pharmacies | 508 | Self-medication | 86% respondents reported had ever practice antibiotic self-medication. Based on health belief model framework, no statistically significant was found in all 4 variables as follows: perceived benefit for not practicing self-medication ($p$=0.989): perceived barrier of no obstacles to practice antibiotic self-medication in terms of drug cost and access to healthcare ($p$=0.928), perception of the threat of antibiotic resistance ($p$=0.232), and the self-efficacy for not practicing antibiotic self-medication ($p$=0.241). |
| **Kondoj**[85] | 2020 | 2019-2020 | Manado, North Sulawesi | Community respondents | 290 | AMR awareness | 43.8% respondents were aware that inappropriate use of antibiotics could lead to AMR. |
| | | | | | | Antibiotic use knowledge | 69% respondents had a low level of knowledge on antibiotic use, 14% moderate level of knowledge, and 17% had good level of knowledge. |

| First author | Year of publication | Year of study | Location | Study population | No. of population | Themes | Summary of findings |
|---|---|---|---|---|---|---|---|
| | | | | buying antibiotics | | Self-medication | Only 20% respondents stated that they always bought antibiotics with prescriptions whereas 12.7% often, 40.6% sometimes, and 26.5% never bought antibiotics with prescriptions. |
| | | | | | | Advice to take antibiotic/ to practice self-medication | 21% stated that they always self-medicated themselves according to advice from friends and relatives. |
| **Kondoj**[85] | 2020 | 2018 | Yogyakarta, DIY | Outpatient at clinics and pharmacies | 268 | AMR awareness | Low awareness of AMR was shown in survey as 80.7% respondents stated that AMR is only a problem for people who take antibiotic regularly, 76.9% said that AMR occurs when the body becomes resistant to antibiotics, and 55.2% were aware that overuse antibiotic can drive AMR. |
| | | | | | | Antibiotic use knowledge | 91% respondent understood that antibiotics are aimed to treat bacterial infections, but 78% stated antibiotics can treat viral infections, and can be used to treat colds and cough (75%). |
| | | | | | | Advice to take antibiotic/ to practice self-medication | 71% respondents agreed that internet is major source of information on antibiotics, while only 58% and 45% consider pharmacists and medical professionals respectively. |
| **Kurniawan**[87] | 2017 | 2015 | Manado, North Sulawesi | Community respondents at primary health centre | 400 | Antibiotic use knowledge | Participants showed good knowledge that antibiotics kill bacteria (73%); moderate knowledge that inappropriate antibiotic use can lead to resistance (59.5%); poor knowledge related to self-medication (67.8%) and the usefulness of antibiotic i.e., for cough or runny nose (74%), does not play a role in viral infection (18%). Participants with lower level of knowledge have higher probabilities to self-medicate with antibiotics than their counterparts. |
| | | | | | | Self-medication | Of all participants, 45% self-medicated in the past six months. |
| | | | | | | Where to buy antibiotic | 52.2% at the pharmacies and 43.9% at kiosks. |
| | | | | | | Advice to take antibiotic/ to practice self-medication | Majority (70.6%) were based on own knowledge, 25,6% from family/friends. The rest from medical workers and pharmacists. |
| | | | | | | Reasons for self-medication | Doctor visit is unpractical (55.6%) or too expensive (3.3%), participants were too busy (26.7%) and used the same antibiotic previously to treat similar symptoms (14.4%). |
| **Novelni**[88] | 2020 | 2019 | Padang, West Sumatra | Community respondents | 100 | AMR awareness | 54% respondents were aware that AMR may lessen the potency of antibiotics to kill the bacteria. |
| | | | | | | Antibiotic use knowledge | 71.2% respondents have moderate-level knowledge on antibiotic use, with the lowest level of knowledge was found in low-education level group. |
| **Nuraini**[89] | 2018 | 2017-2018 | Bangkalan, East Java | Outpatients at secondary hospital | 103 | Antibiotic use knowledge | Based on health belief model framework, the perceived benefit of antibiotic use ($p$=0.021) was statistically significant, while perceived barrier to antibiotic use ($p$=0.426), perceived threat to antibiotic use ($p$=0.167), and self-efficacy of antibiotic treatment ($p$=0.872) were not statistically significant. |

| First author | Year of publication | Year of study | Location | Study population | No. of population | Themes | Summary of findings |
|---|---|---|---|---|---|---|---|
| | | | | | | Adherence to treatment | Based on leftover pill counting, 55.3% completed the full antibiotic course and 44.7% did not. Participants stopped the antibiotic course because they felt better and fear of side-effects on the renal function. |
| **Pratama**[90] | 2018 | 2016 | Jember, East Java | Community respondents: non-medical university students | 324 | Self-medication | 68.5% ever used antibiotics of which 58.6% reported to purchase antibiotic without prescription. Those who have insurance were less likely to self-medicate (50%) compared to those who did not have an insurance (64%). |
| | | | | | | Where to buy antibiotic | 46.3% at the pharmacy, 12.9% at the grocery store, 12.4% from family/friends, and 10.1% at drug store. |
| **Salsabila**[91] | 2020 | 2019 | Yogyakarta, DIY | Community respondents in rural and urban areas | 125 | AMR awareness | 76.8% respondents were aware that bacteria could be resistant to antibiotics. However, 53.6% had low level of overall knowledge on antibiotic resistance. |
| | | | | | | Antibiotic use knowledge | 53.6% respondents had low level of on antibiotics use and resistance, while 46.4% had high level of knowledge. |
| | | | | | | Advice to take antibiotic/ to practice self-medication | 78.4% respondents sought healthcare professional for information on antibiotics, while 21.6% consulted internet and social media. |
| **Siahaan**[92] | 2017 | 2015 | No city mentioned, West Java, DKI Jakarta, Southeast Sulawesi | Community respondents: household members | 1271 | Adherence to treatment | 66.5% of participants took antibiotic according to doctor's instructions, and 33.9% took it only until they would feel better. |
| **Tandjung**[94] | 2021 | 2020-2021 | Manado, North Sulawesi | Community respondents | 323 | Antibiotic use knowledge | 50.5% respondents had low level of knowledge based on the response on 13 questions. 21% practiced irrational antibiotic use based on 6 indicators: information about how to use antibiotics, indication of antibiotic use, duration, dose, follow up, and dispensing. |
| | | | | | | Self-medication | 80% respondents reported ever bought antibiotics without prescriptions opposed to the rest of 20% respondents. |
| | | | | | | Reasons for self-medication | Previous use (82%), cost-effectiveness (72%), previous similar symptoms (80%). Pharmacists allowed antibiotic purchase without prescriptions (71%). |
| | | | | | | Advice to take antibiotic/ to practice self-medication | 78% respondents used unprescribed antibiotics suggested by friends or relatives. |
| **WHO**[95] | 2015 | 2015 | National | Community respondents: | 1027 | AMR awareness | 84% were familiar with the term AMR, 68% considered antibiotic resistance is one of the biggest world problems and 67% believed that many infections are becoming resistant to antibiotics. |

| First author | Year of publication | Year of study | Location | Study population | No. of population | Themes | Summary of findings |
|---|---|---|---|---|---|---|---|
| | | | | household members | | Antibiotic use knowledge | 66% knew what type of disease that can be treated with antibiotic, but 63% thought cold and flu could be treated with antibiotic. 76% knew they have to take the full course of antibiotic treatment. 86% thought people should use antibiotics only when they are prescribed by doctors, but 51% thought it is ok to buy the same antibiotic if it helped previously. However, 64% disapproved of using an antibiotic given by family/friend even it was aimed to treat the same illness. The majority took antibiotics in the past month (30%) and past six months (36%). In the last doctor's visit, 83% were prescribed with antibiotics; 87% got advice on how to take antibiotic. |
| | | | | | | Where to buy antibiotic | 96% at the pharmacy, 1% at stall or hawker, 1% from the internet, 1% saved up from previous time. |
| | | | | | | Advice to take antibiotic/ to practice self-medication | 83% from doctors/nurses, 15% did not get advice from doctors/nurses, and 1% could now remember. |
| | | | | | | Antibiotic stewardship | 88% stated that doctors should only prescribe antibiotics when needed, and 83% thought governments should promote the development of new antibiotics. However, the majority (75%) thought that lay people cannot do anything to stop AMR and believed that medical experts will solve the problem before it becomes too serious |
| **Widayati**[96] | 2011 | 2010 | Yogyakarta, DIY | Community respondents: household members | 559 | Self-medication | Of all participants who self-medicated, 80% had previous experience in using such antibiotics. A positive association was found between self-medication and having health insurance. |
| | | | | | | Where to buy antibiotic | 64% at the pharmacy, 19.5% at both pharmacy, drug store, and kiosk. |
| | | | | | | Advice to take antibiotic/ to practice self-medication | Physician or pharmacist (51.2%); friends/relatives (31.7%); multiple sources (9.8%); magazine, advertising, and brochure (4.9%); and past experiences (2.4%). |
| | | | | | | Reasons for self-medication | Reasons for self-medication were previous positive experience (54%), saving time and saving money. |
| **Widayati**[97] | 2012 | 2010 | Yogyakarta, DIY | Community respondents: household members | 559 | AMR awareness | 85% have appropriate knowledge regarding antibiotic resistance, 76% about antibiotic use for bacterial infections, but 71% had incorrect knowledge regarding antibiotics for viral infections. 50% knew that antibiotics not to be used immediately for fever. Good knowledge correlated with being male, young, higher education, and higher income. |
| | | | | | | Antibiotic use knowledge | 74% believe that antibiotics can prevent illnesses from becoming worse and 40% believe antibiotic can cure any illness. |

| First author | Year of publication | Year of study | Location | Study population | No. of population | Themes | Summary of findings |
|---|---|---|---|---|---|---|---|
| **Yulia**[98] | 2019 | 2019 | Bukittinggi, West Sumatra | Community respondents | 100 | AMR awareness | 65% respondents were aware that AMR may lessen the potency of antibiotics to kill the bacteria, while 35% respondents were not. 68% did not aware that when pathogens are resistant to one antibiotic, other antibiotics might be potent to kill the pathogen. |
| | | | | | | Antibiotic use knowledge | 40% respondents stated antibiotic can cure all disease and 67% stated antibiotic can cure fungal infection. 80% were aware that full antibiotic course must be completed and 99% aware that antibiotic must be used based on doctor's instruction. |
| **Yuliani**[99] | 2014 | 2014 | Kupang, East Nusa Tenggara | Community respondents | 100 | Antibiotic use knowledge | Important misconceptions: 94% thought antibiotics can treat every disease, 92% thought that every medicine is in the antibiotic group, 94% thought that every disease can be treated with antibiotics and 68% think that non-infectious disease, such as cold, fever, headache can be treated with antibiotics. |
| **Health care providers** | | | | | | | |
| **Asvinigita**[77] | 2019 | 2019 | Yogyakarta, DIY | Healthcare providers: community pharmacists | 250 | Antibiotic dispensing | Only 76% of pharmacists would refer the patients to physicians when symptoms are suggestive of an infection. 68% stated they would dispense antibiotic without prescriptions. Respondents had generally positive view on antimicrobial stewardship programme |
| **Siswati**[93] | 2009 | 2000 | Padang, West Sumatra | Healthcare providers | 110 | Antibiotic dispensing | 98.8% did not respond to patients' antibiotic demand, but 12.1% gave antibiotic so the patient could leave quickly. Advice to prescribe antibiotics were obtained from community health centre doctor (33.3%), paediatricians (17.2%), and the rests were unspecified. Factors that were associated with antibiotic prescribing were level of antibiotic knowledge, attitude toward clinical guidelines, and supervision. |
| | | | | | | Antibiotic use knowledge | 54.5% participants had high knowledge and 45.5% had low knowledge. Most respondents (77.9%) never received training on rational drug use. Only 44% had other health care providers as role models in using antibiotics, while 55.6% did not. |
| | | | | | | The use of antibiotic guideline | 63.6% showed a positive attitude towards treatment guidelines, 35.4% showed a slightly positive attitude, 1% showed a slight negative attitude, and no one showed a negative attitude. |
| **Zhang**[100] | 2020 | 2018 | National | Healthcare providers (General practitioners and paediatricians) | 100 | Antibiotic dispensing | Physicians stated that around 47% medical consultation led to antibiotic dispensing, with the most common age-group to receive antibiotics was 0-1year old. Physicians in Indonesia considered side effects of antibiotics as the highest consideration that detracted them from prescribing antibiotics to children. Only 19% respondents stated they would prescribe antibiotics to a 4-yo child with a 2-day history of upper respiratory tract infection. The major determinant to prescribing antibiotics was the severity of specific clinical symptoms. |

Abbreviations: AMR, antimicrobial resistance; DIY, Daerah Istimewa Yogyakarta; DKI, Daerah Khusus Ibukota; WHO, World Health Organization
